# Spatio-temporal associations between deforestation and malaria incidence in Lao PDR

**DOI:** 10.1101/2020.04.23.20072215

**Authors:** Francois Rerolle, Emily Dantzer, Andrew A. Lover, John M. Marshall, Bouasy Hongvanthong, Hugh JW. Sturrock, Adam Bennett

## Abstract

As countries in the Greater Mekong Sub-region (GMS) increasingly focus their malaria control and elimination efforts on forest-going populations, greater understanding of the relationship between deforestation and malaria incidence will be essential for programs to assess and meet their 2030 elimination goals. Leveraging village-level health facility surveillance data and forest cover data in a spatio-temporal modeling framework, we found evidence that deforestation is associated with short-term increases, but long-term decreases in confirmed malaria case incidence in Lao People’s Democratic Republic (Lao PDR). We identified strong associations with deforestation measured within 30 km of villages but not with deforestation in the near (10 km) and immediate (1 km) vicinity. Results appear driven by deforestation in densely forested areas and were more pronounced for infections with *Plasmodium falciparum* (*P. falciparum*) than for *Plasmodium vivax* (*P. vivax*). These findings highlight the influence of forest-going populations on malaria transmission in the GMS.

## Introduction

Engaging in forest activities, such as logging, hunting or spending the night in the forest, is considered a primary risk factor for malaria infection in the Greater Mekong Sub-region (GMS) [17, 20, 39, 22, 61] with recent outbreaks attributed to deforestation activities [42]. This is most likely the result of increased human exposure to the forest dwelling *Anopheles dirus* and *Anopheles minimus*, the most efficient and widespread malaria vectors in the GMS [47, 46]. However, deforestation may also alter this “forest malaria” [65] ecosystem and eventually reduce malaria incidence, as is generally accepted to be the case in Southeast Asia [29]. Several previous studies have assessed the relationship between deforestation and malaria, but the majority focused outside of the GMS, most notably in the Amazonian forest [67, 48, 31, 64, 60] where the evidence has been met with conflicting interpretations [62]. As national malaria programs across the GMS target forest-going populations for prevention and treatment efforts [30, 57], improved understanding of the relationship between deforestation and malaria is critical for programs to assess and meet national 2030 malaria elimination goals [49, 8].

In the Amazon, the “frontier malaria” hypothesis [54] posits that malaria temporarily increases following deforestation efforts to open a human settlement area in the forest. Subsequently, after approximately 6-8 years, settlements become more developed and isolated from the surrounding forest, and less suitable for malaria vectors, resulting in reduced malaria transmission [16]. Recent work has challenged this hypothesis, however, and found that some older settlements were also likely to have high malaria incidence [36], highlighting the importance of assessing the relationship between deforestation and malaria at different spatio-temporal scales [55]. Similarly, a recent review of the literature on deforestation and malaria in the Amazon [62] highlights the inconclusive evidence on the complex links between forests, deforestation and malaria.

The importance of addressing complex confounding structures influencing the relationship between deforestation and malaria was highlighted by Bauhoff et al. [12]. Variables such as temperature [13, 44], precipitation [51, 50] or seasonality [35] are known environmental predictors of malaria, although the spatio-temporal scale of those effects if often highly variable across different areas [59]. Furthermore, remote areas may experience higher malaria rates because of poor access to public health services, but also have denser forest cover or lower deforestation rates [15]. Finally, forest-going populations in the GMS are also at higher risk for malaria [52] due to poor adherence to protective measures against mosquitoes such as insecticide-treated bed nets (ITNs) or long-lasting insecticidal hammocks (LLIHs) [28, 57] and delayed and inadequate access to treatment [30].

Bauhoff et al. [12] identified only 10 empirical studies that assessed the relationship between deforestation and malaria with appropriate adjustments for confounding. Of these, seven reported a positive association [10, 67, 48, 60, 53, 27, 25], two did not find any associations [12, 31], and one disputed study found a negative association [64, 32, 63]. Most recently, a study found deforestation to increase malaria risk and malaria to decrease deforestation activities in the Amazon, using an instrumental variable analysis to disentangle any reverse causality loop [43]. However, only half of the above-mentioned studies used high-resolution forest data, with most studies using spatially aggregated data and exploring only a limited range of spatial and temporal scales. Only three of these studies were conducted in Southeast Asia [53, 27, 25], and none in the GMS. Importantly, all three found that malaria increases after deforestation, but all had limitations. The two studies in Indonesia [53, 27] used coarsely aggregated forest data and potentially biased self-reported malaria data. The third study, in Malaysia [25], focused on a specific and geographically confined malaria parasite, *Plasmodium knowlesi*, whose primary host is the long-tailed macaque and whose presence in the GMS, where *P. falciparum* and *P. vivax* dominate, is limited.

In this paper, we used Landsat 30 m high-resolution forest data [34] and monthly malaria incidence data from two separate regions in the GMS: northern Lao People’s Democratic Republic (PDR) which is approaching malaria elimination and southern Lao PDR where *P. falciparum* and *P*. *vivax* are co-endemic. By conducting the analysis at the village level, we were able to explore a wide range of spatial scales (1, 10 and 30 km around villages) that might be relevant in characterizing the relationship between deforestation and malaria. In addition, we leveraged the longitudinal nature of both the incidence data collected and the forest data produced from annual remote sensing imagery [34] to explore the most relevant temporal scales. Finally, we considered alternative definitions of deforestation, restricted to areas with at least certain levels of forest cover, to investigate the type of deforestation driving the relationship with malaria.

To date, no prior studies have quantified the relationship between deforestation and malaria incidence in the GMS. Understanding this relationship is especially important in the GMS, where forest-going activities are a main source of income generation [6] and malaria clusters in forest-going populations [49, 57]. To assess the relationship between deforestation and malaria incidence, we modeled the monthly village-level malaria incidence in two regions of Lao PDR using health facility surveillance data and evaluated the most relevant spatio-temporal scale.

## Results

### Forest and environmental variables

Figure 1 shows the average tree crown cover within 10 km for the year 2016 and the percent area within 10 km that experienced forest loss between 2011 and 2016 in 2 regions of southern and northern Lao PDR. Overall, the forest cover was denser in the north than in the south and deforestation over this period was higher in the north than in the south. Figures 11 and 12 in the appendix show the distribution of forest and deforestation variables as the temporal scales and spatial scales around study villages were varied. For example, within 30 km of a village, the percent area that experienced forest loss between 2011 and 2016 in the north ranged between 0 and 10% whereas it rarely exceeded 2.5% in the south. As expected, the longer the time period considered, the larger the areas that experienced forest loss. The average forest cover increased with increasing buffer radius around villages (1, 10 and 30 km). However, the relationship with the percent area that experienced forest loss was less clear because both the area that experienced forest loss (numerator) and the area around villages (denominator) increased.

**Figure 1.**
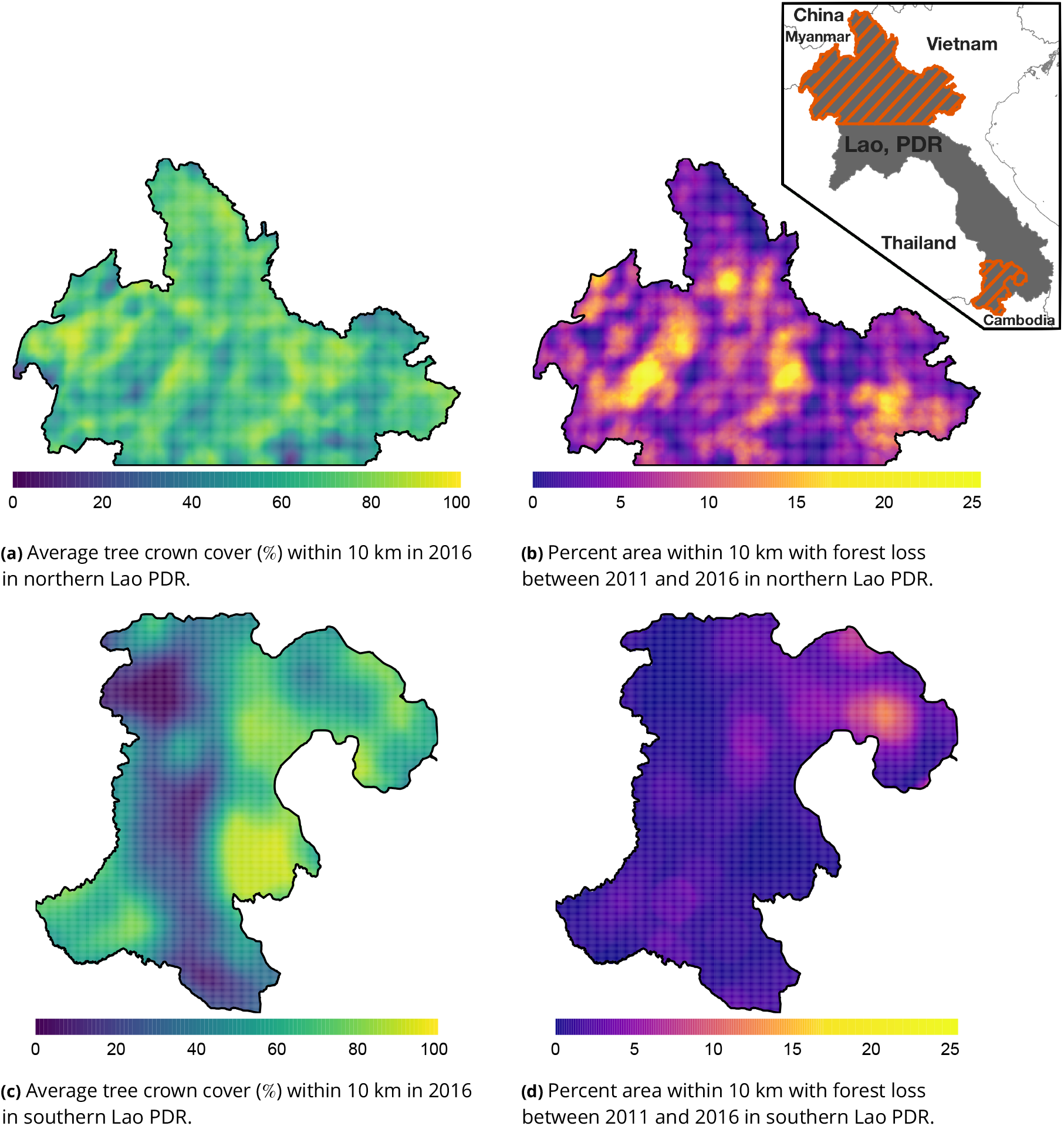
Average tree crown cover (%) in 2016 (left) and percent area that experienced forest loss between 2011 and 2016 (right) within 10 km in northern (top) and southern (bottom) Lao PDR. Upper right indent maps northern and southern (Champasak province) Lao PDR regions.

**Figure 2.**
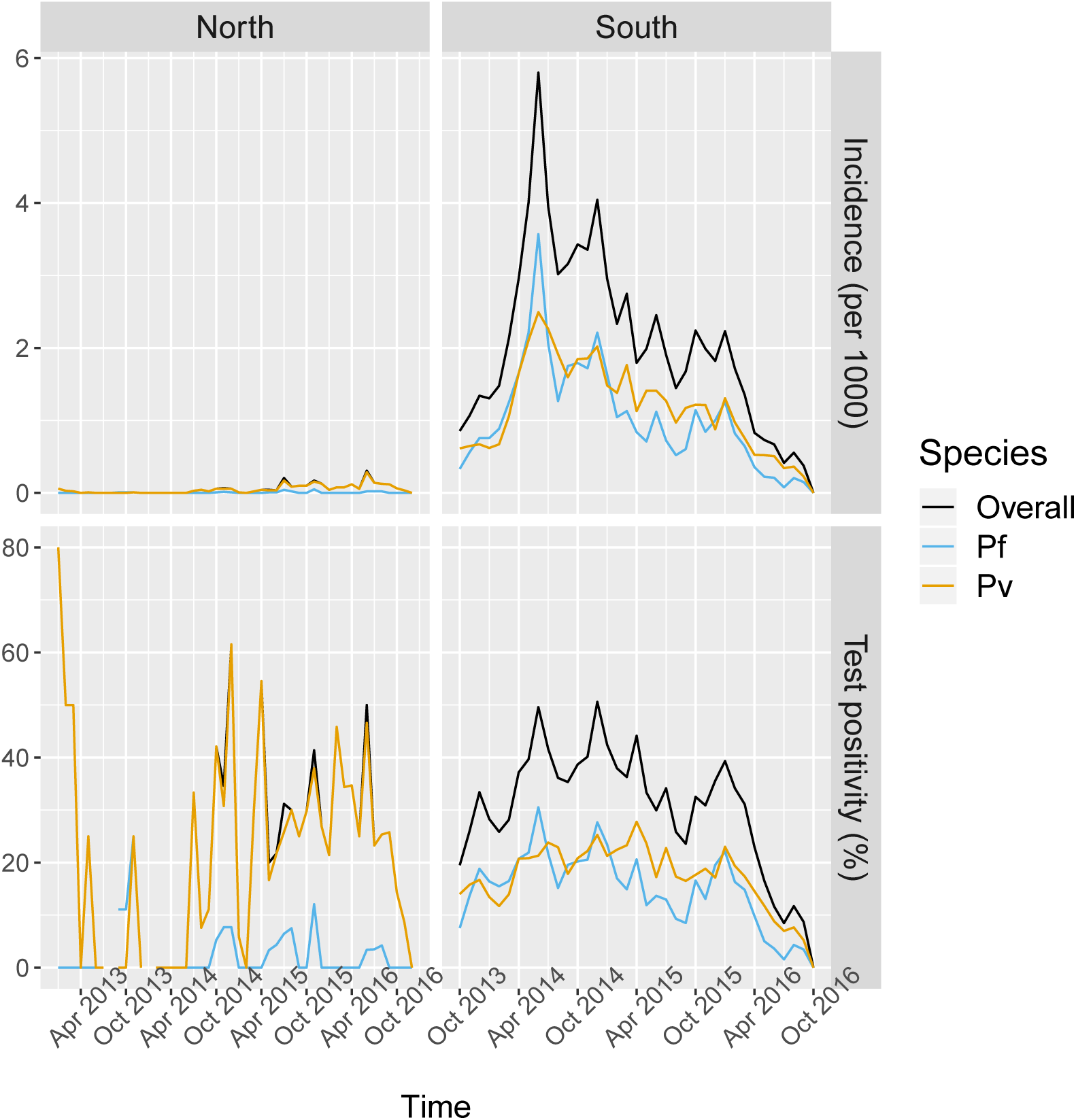
Malaria incidence (per 1000) and test positivity (%) over time.

**Figure 3.**
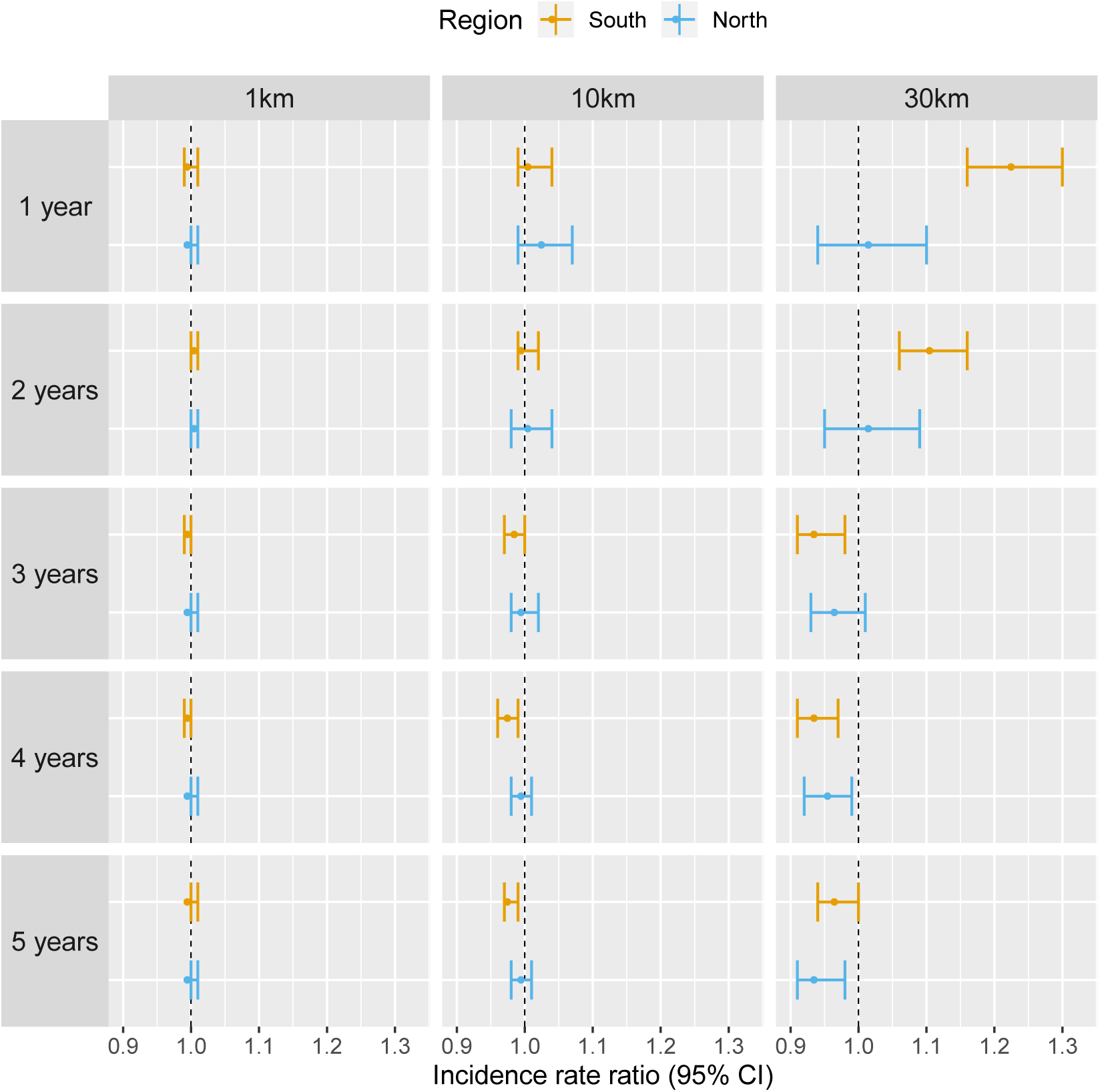
Associations between malaria incidence and a 0.1 % increase in the area that experienced deforestation within 1,10 or 30 km (left-right) of a village in the previous 1 to 5 years (top-down) in Lao PDR. Adjusted for the spatio-temporal structure of the data, the environmental covariates selected in the model and forest cover within 30 km in the year before the deforestation temporal scale considered.

**Figure 4.**
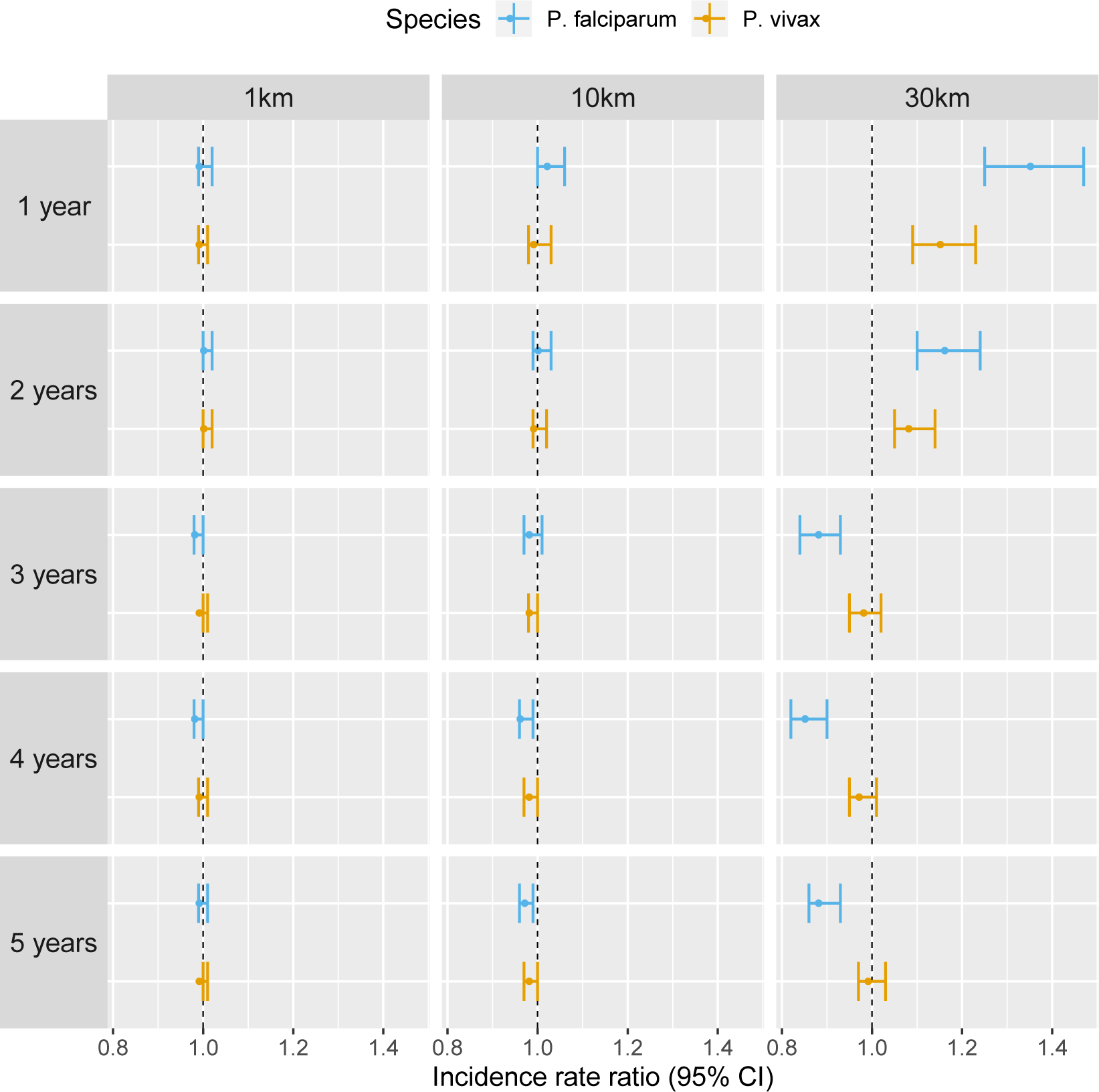
Associations between malaria incidence and a 0.1 % increase in the area that experienced deforestation within 1,10 or 30 km (left-right) of a village in the previous 1 to 5 years (top-down) in southern Lao PDR, differentiated by malaria species. Adjusted for the spatio-temporal structure of the data, the environmental covariates selected in the model and forest cover within 30 km in the year before the deforestation temporal scale considered.

**Figure 5.**
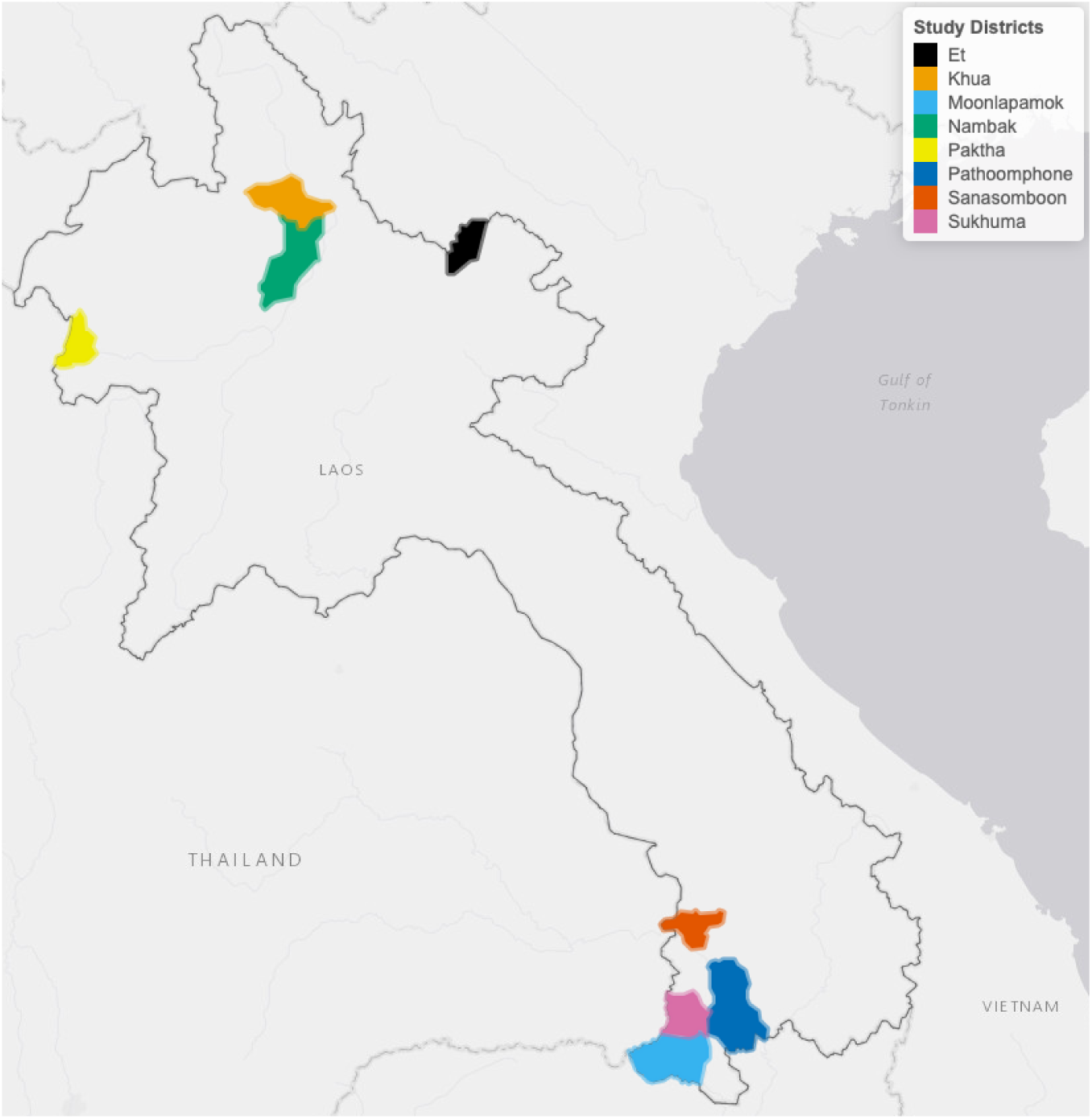
Map of study’s districts

**Figure 6.**
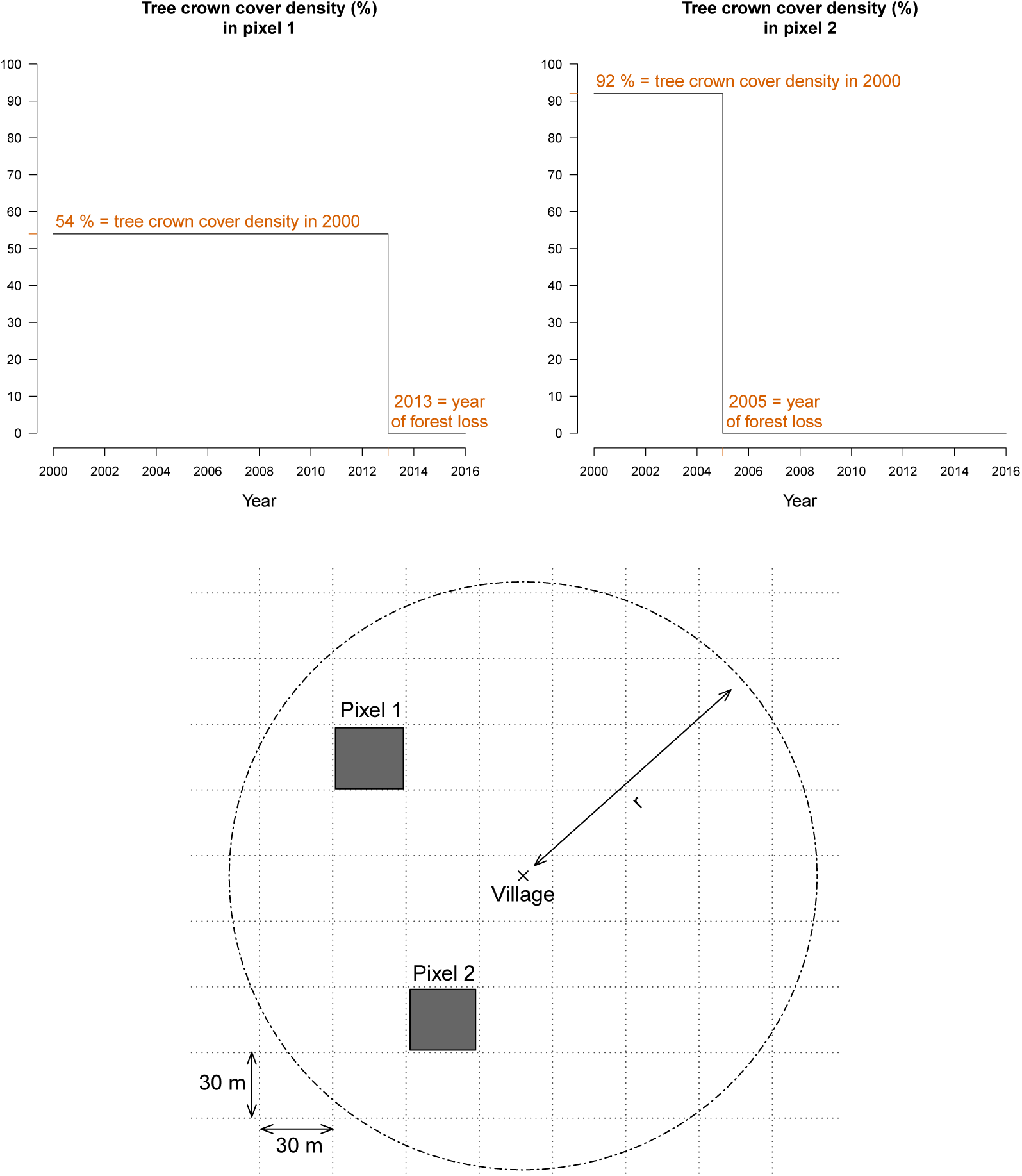
For every 30m Landsat pixel within a buffer radius r(1,10 and 30 km) ofstudy’s villages, the tree crown cover density in 2000 and the year of forest loss were combined to derive the deforestation and forest cover variables. The two upper plots highlight the raw data at two example pixels from the lower plot.

**Figure 7.**
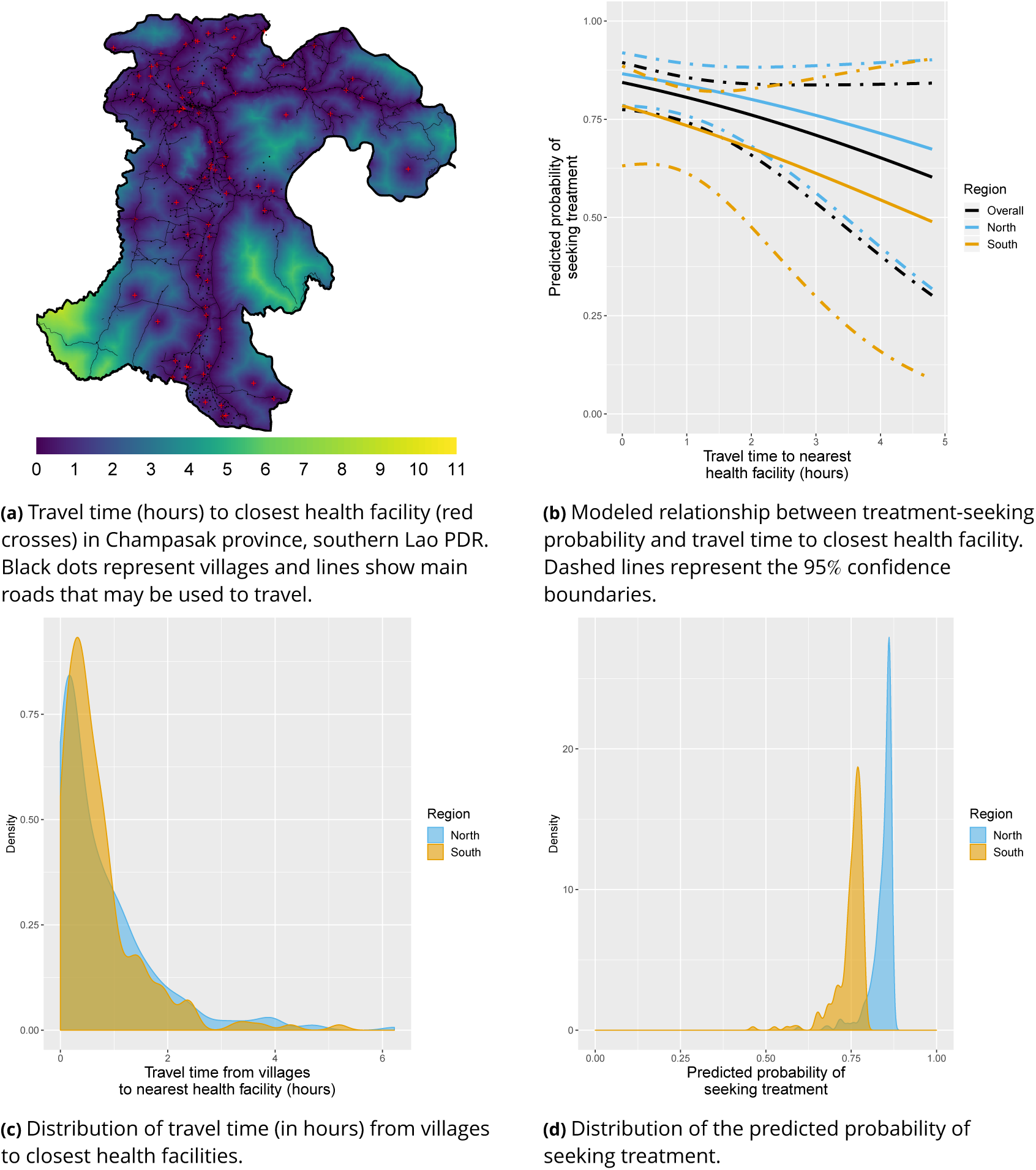
Treatment-seeking modeling plots. Note that treatment-seeking at public health facilities is implied all along the manuscript.

**Figure 8.**
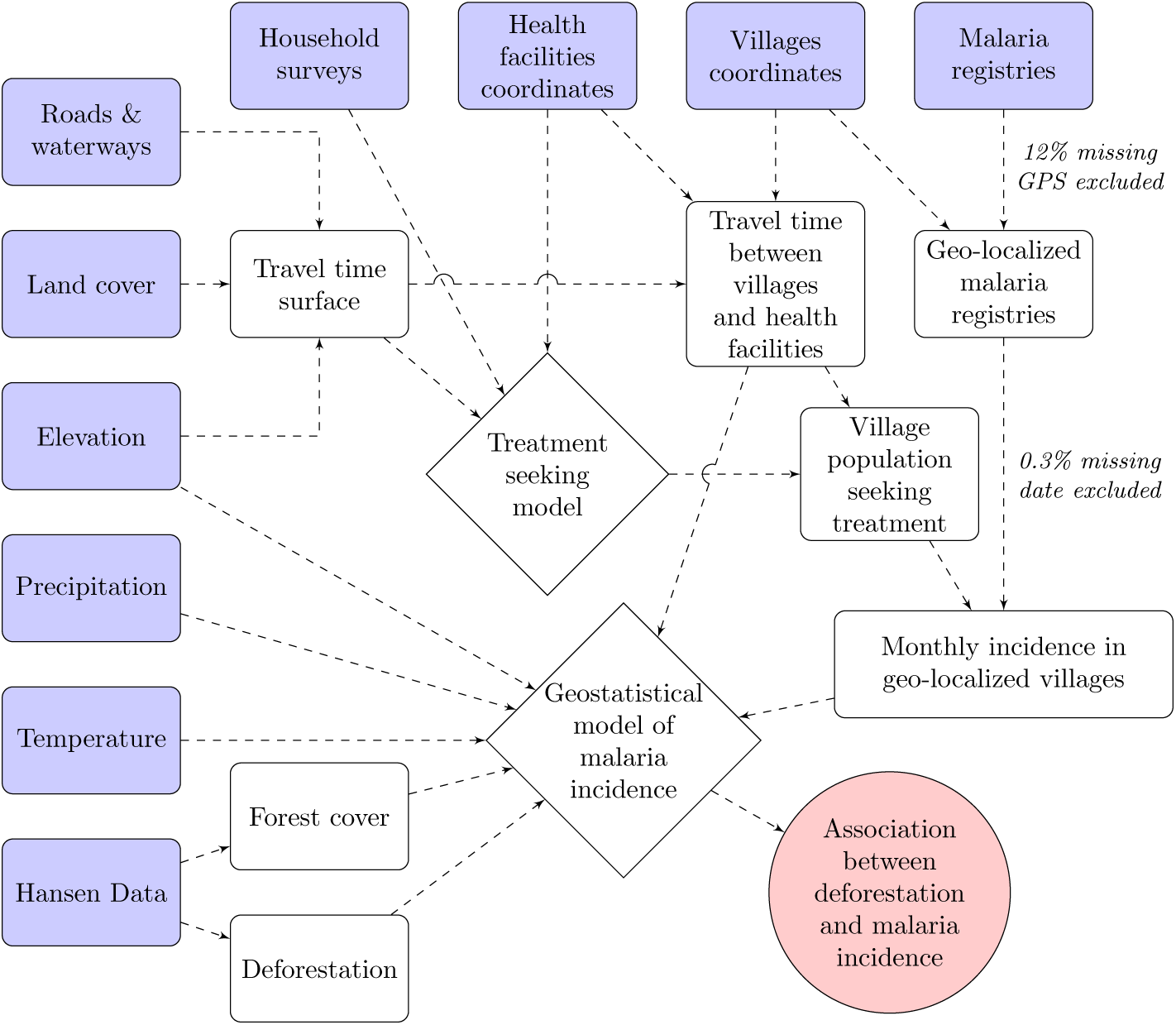
Conceptual model for our analysis showing how the raw input data (blue boxes) were combined via intermediate data (white boxes) and models (white diamonds) to produce our estimated outputs (red circle).

**Figure 9.**
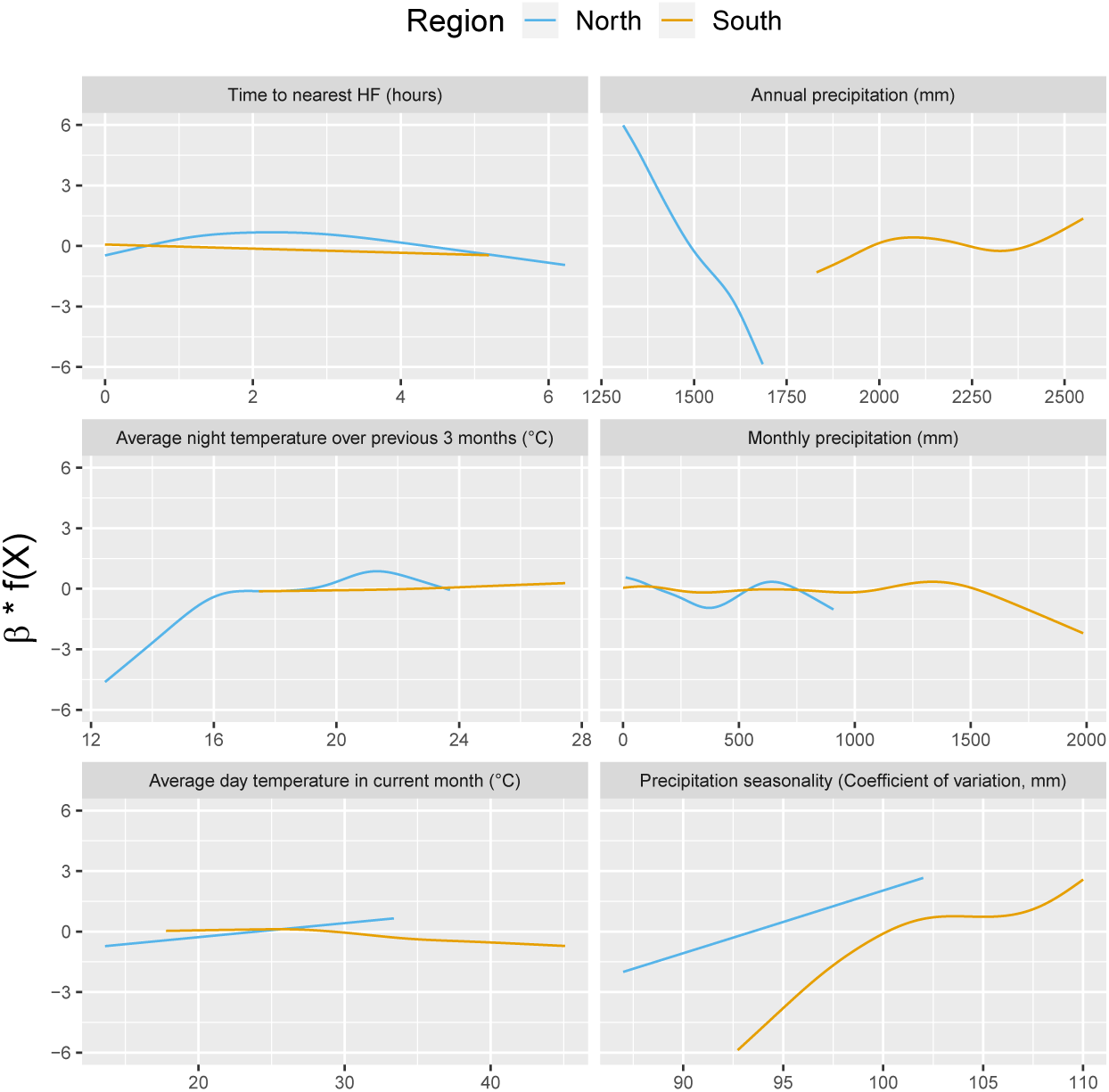
Relationships between malaria incidence and environmental covariates in the multivariable model selected by the forward selection approach, as described in the methods section. The model was additionally adjusted for the spatio-temporal structure of the data (*f* (*t*), *f* (*Lat, Long*) and village random intercepts). Note that 95% confidence intervals (see Fig. 18 in the appendix) have been hidden for better visualization.

**Figure 10.**
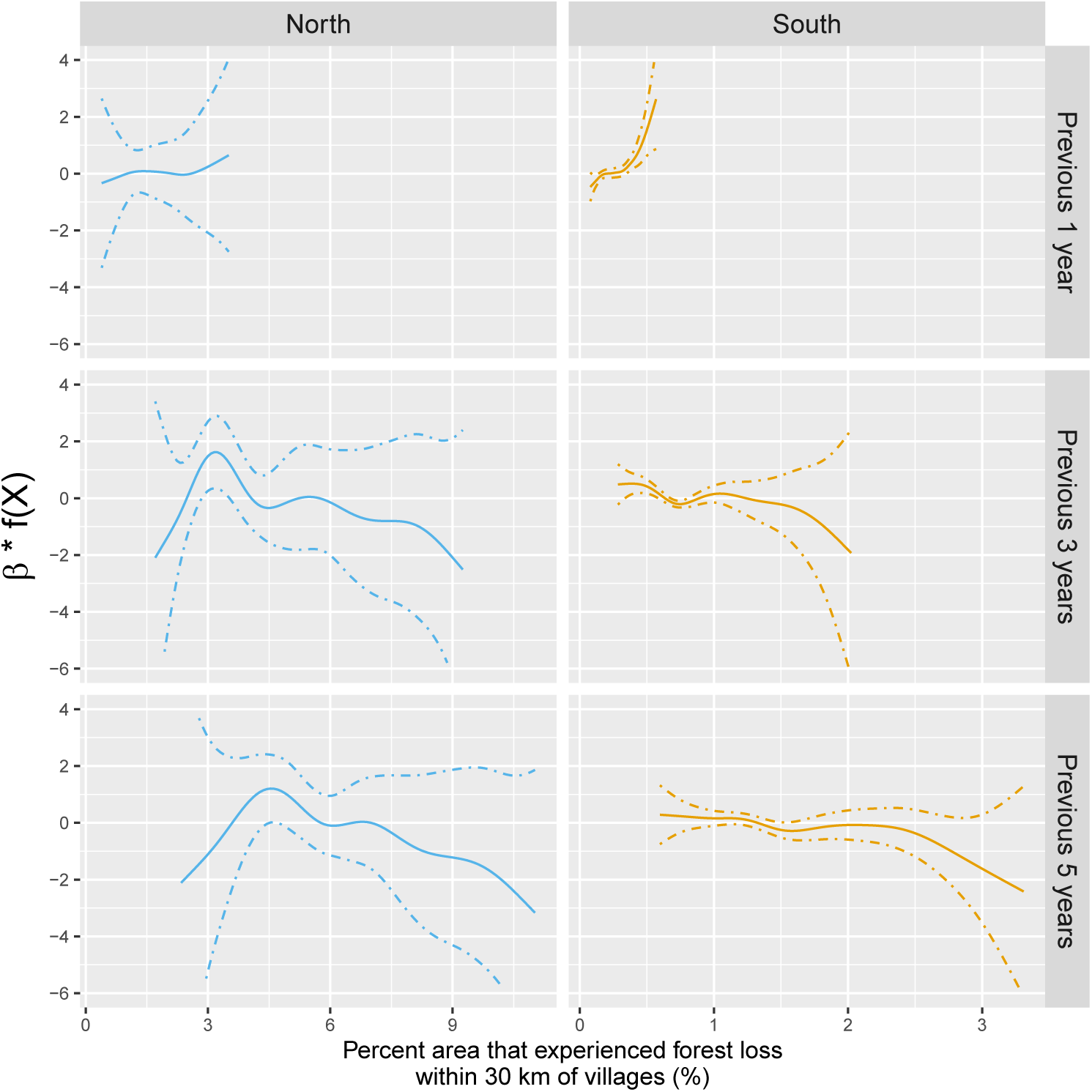
Adjusted relationship between deforestation and malaria incidence. All models were adjusted for environmental covariates and forest cover selected by the model selection approach as described in the methods section on top of the spatio-temporal structure ofthe data (*f*(*t*), *f* (*Lat, Long*) and village random intercepts). Note that scales are different between regions for better visualization. Figure 19 in the appendix showsthe raw scatterplot between monthly village malaria incidence rate and deforestation. Figures 20 and 21 in the appendix show the raw time series of malaria incidence, forest cover and percent area that experienced forest loss.

**Figure 11.**
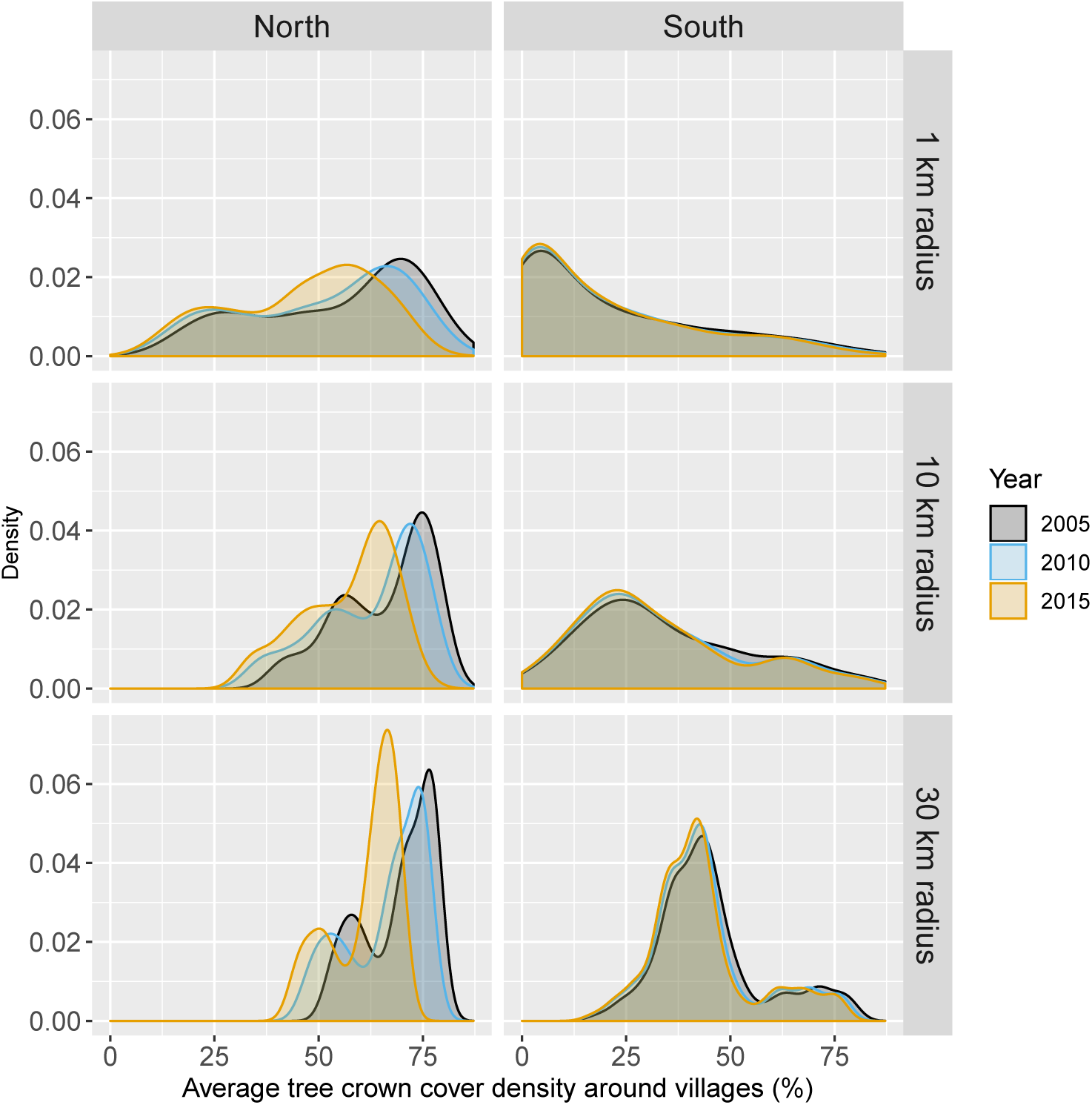
Distribution of average tree crown cover density within 1,10 and 30 km of villages.

**Figure 12.**
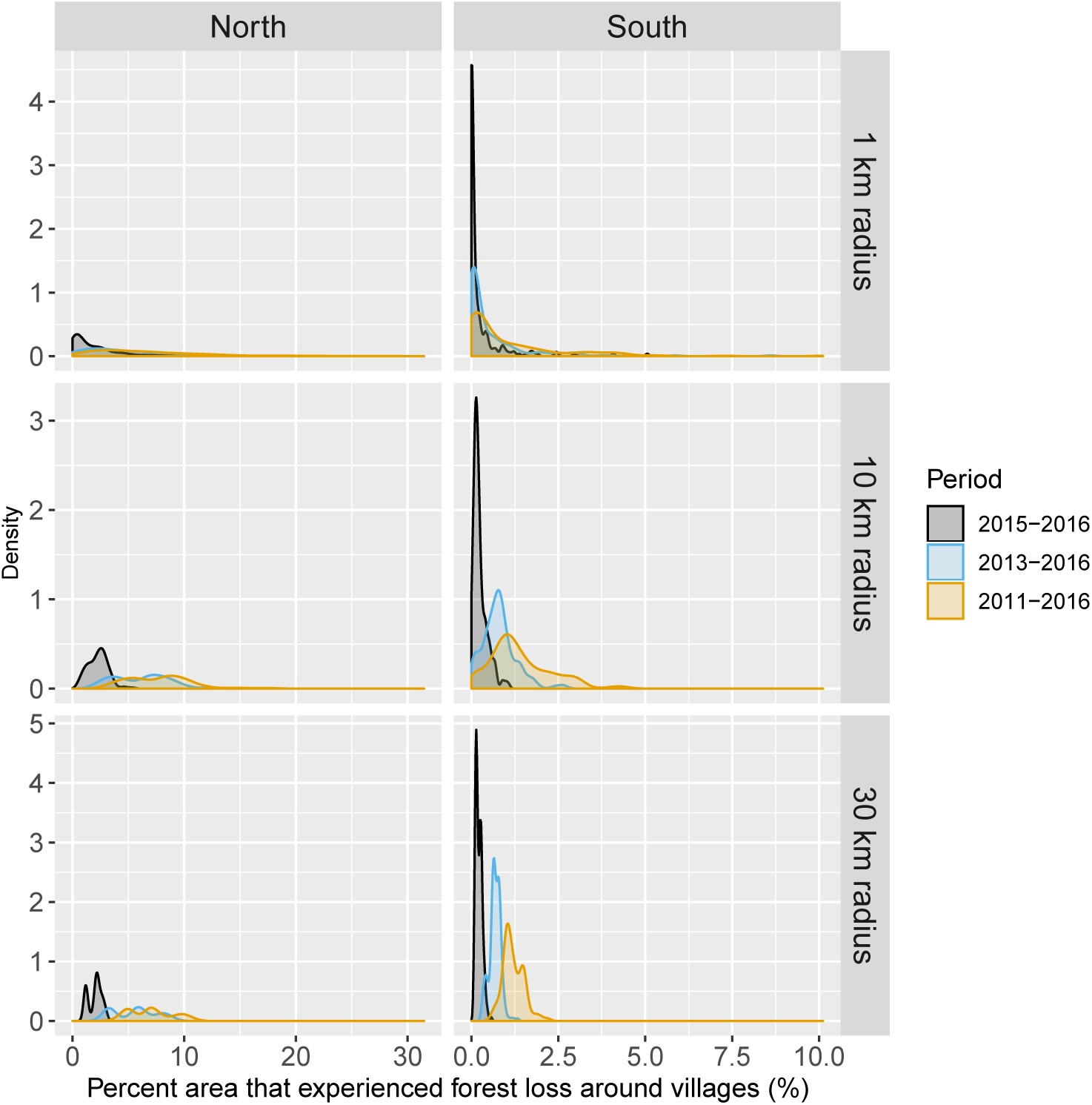
Distribution of percent area within 1, 10 and 30 km of villages that experienced forest loss between 2011 and 2016. Note that the scales are different for every panel for better visualization of the distributions.

The 284 villages in the north were overall less populated (mean population size: 498, IQR: [241; 548]) than the 207 villages in the south (mean: 1095, IQR: [584; 1384]), but with some highly populated outliers. As expected, altitude differed substantially between villages of the mountainous northern region (mean: 557m, IQR: [378; 679]) and the lowlands of the south (mean: 120m, IQR: [98; 130]). Although both regions exhibited similar seasonal trends in precipitation and temperature, with a rainy season spanning from April to October, villages in the south experienced higher monthly precipitation and temperature than in the north over the study period (Fig. 13 in appendix).

**Figure 13.**
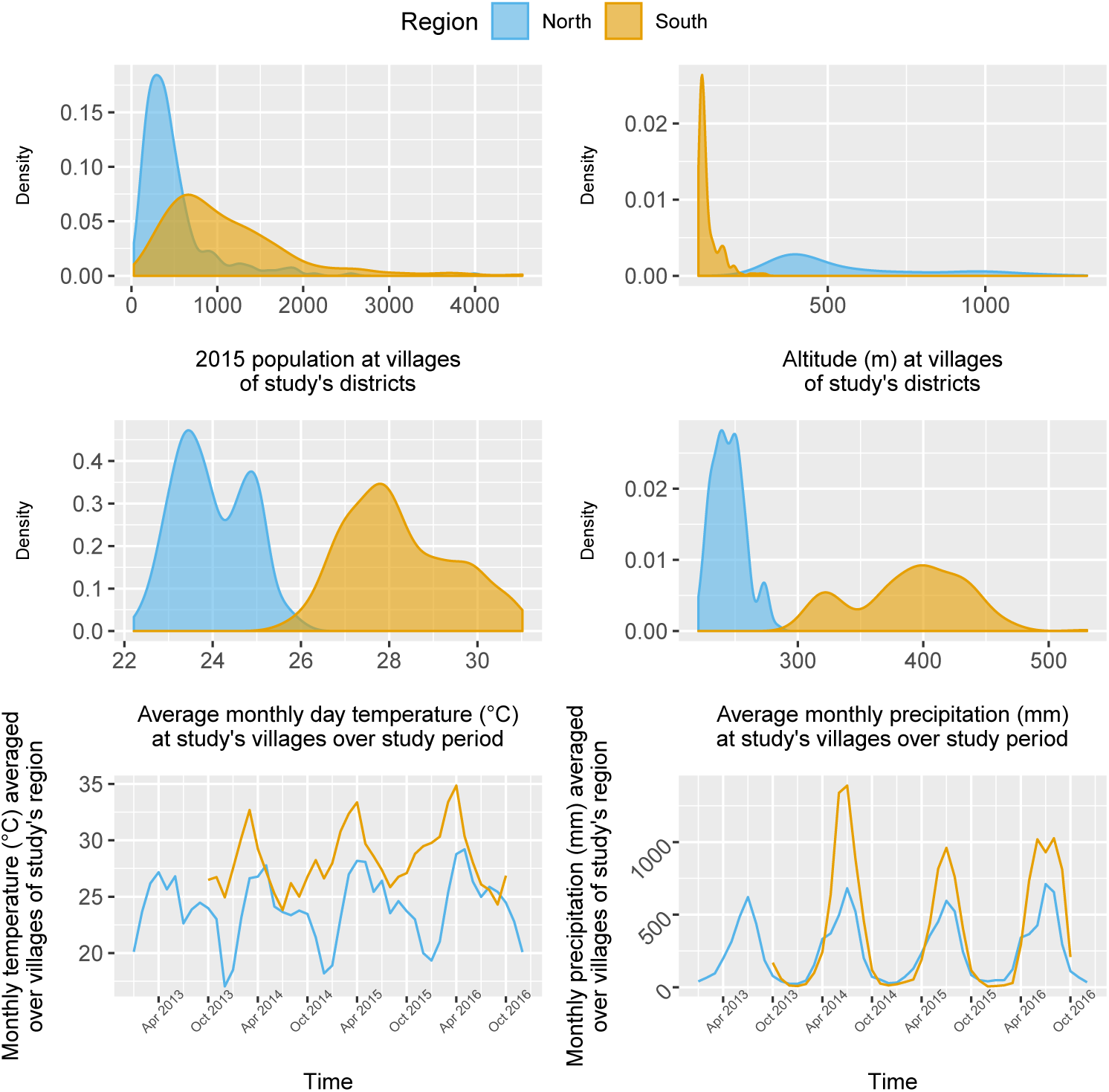
Distribution and time series of environmental covariates (population, altitude, monthly day temperature and monthly total precipitation) at study’s villages.

### Malaria case data

#### Malaria infections

63,040 patient records were abstracted from the malaria registries of all public health facilities in 4 southern districts between October 2013 and October 2016 and 1,754 from all health facilities in 4 northern districts between January 2013 and December 2016.

In the south, 91.2% of the patients in the registries were tested for malaria, of which 78.1 % were tested by RDT and 26.2% by microscopy. Overall test positivity was 33.2% for any infection, 16.4% for *P. falciparum* and 18.2% for *P. vivax*. Monthly incidence peaked to about 6 cases per 1000 people in the 2014 rainy season, eventually decreasing to below 1 case per 1000 in 2016. Incidence and test positivity were similar between *P. falciparum* and *P. vivax* in the south (Fig. 2).

In the north, 92.1 % of the patients in the registries were tested for malaria, of which 96.3% were tested by RDT and 9.6% by microscopy. Overall test positivity was 23.8% for any infection, 2.8% for *P. falciparum* and 22.5% for *P. vivax*. Monthly malaria incidence in the north was very low, never exceeding 0.3 per 1000 people. Most infections in the north were *P. vivax* cases with only a few seasonal *P. falciparum* cases (Fig. 2).

In the appendix, Figure 14 shows the number of patients and cases recorded per month in health facility malaria registries as well as how the smoothed test positivity rates varied over time.

**Figure 14.**
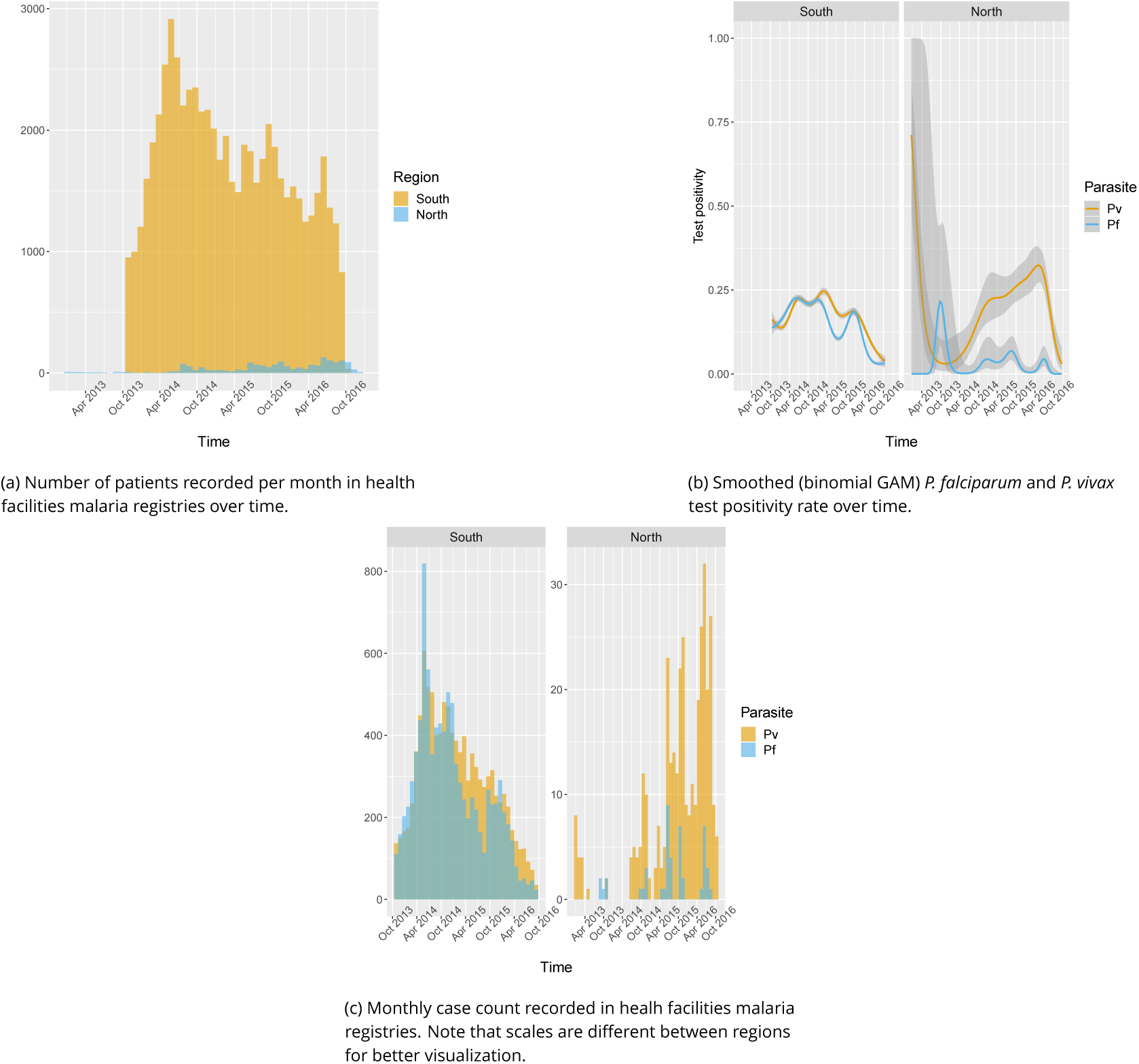
Additional figures from malaria registries: malaria infections

#### Socio-demographics

Age, gender and occupation of patients seeking treatment at health facilities were also recorded in the malaria registries. On average, patients in the south were older than patients in the north with mean age of 28 years and 23 years respectively. In the north, about half of the patients were male (53.1 %), while most patients in the south were male (71.1 %). Finally, the vast majority (68.2%) of patients in the south were farmers, whereas only 8% of patients in the north were farmers. Most patients in the north reported being unemployed (41.7%) or a student (31.2%) (Fig. 15 in the appendix).

**Figure 15.**
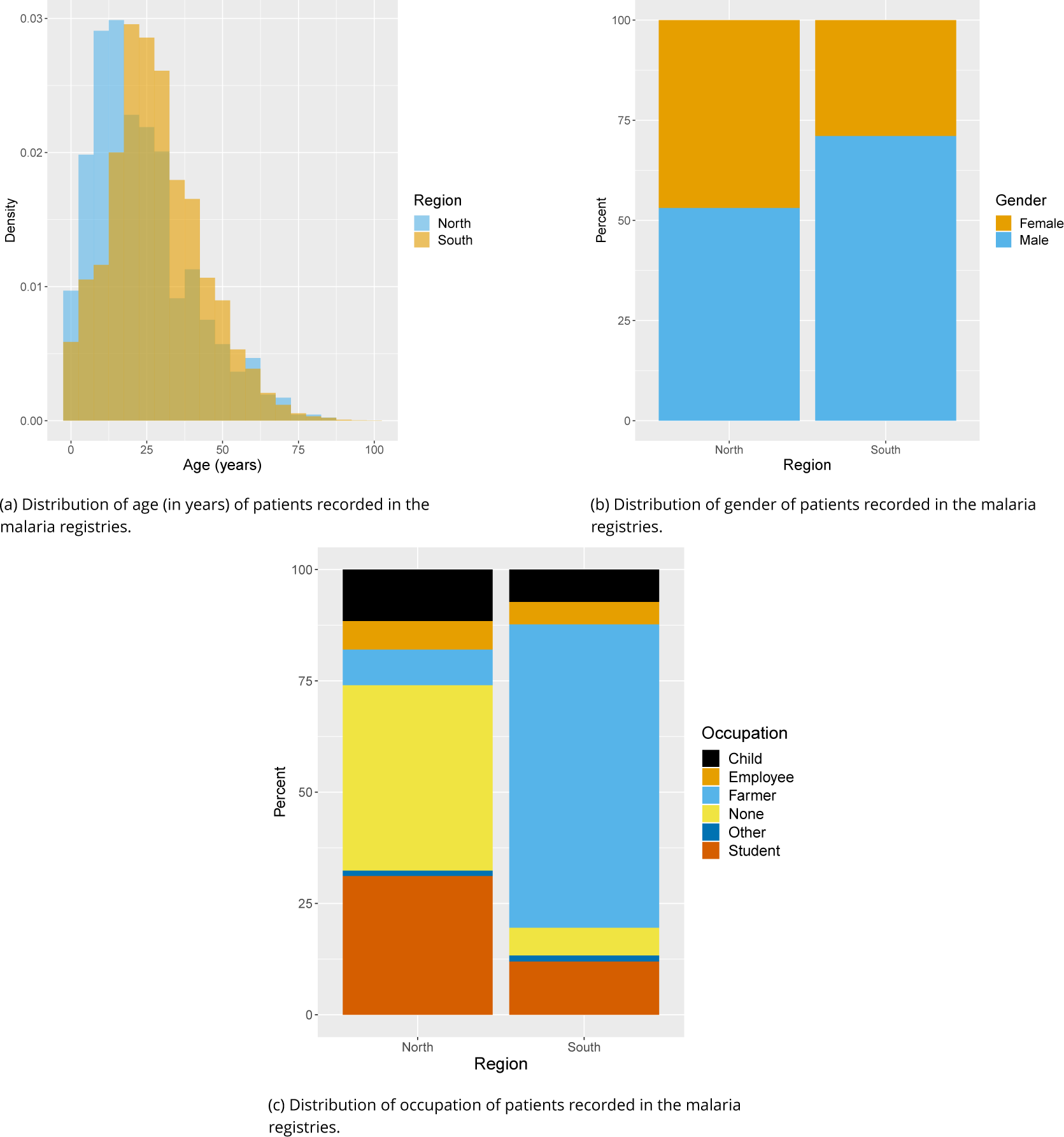
Distributions of socio-economomical variables of all patients recorded in the malaria registries

#### Geo-referencing

Overall, 88.1 % of malaria records were matched to one of the 491 villages in study districts. The remaining (11.7% in the south and 17.3% in the north) were removed from the analysis because of ambiguous village names, local nicknames for small villages and dissolving and grouping of villages over time. Test positivity in the south was similar in matched (33.1%) and unmatched (34.2%) records but higher in matched (26.5%) than unmatched (10.5%) records in the north. No substantial difference was found in the distribution of socio-demographic variables available in malaria registries between matched and unmatched records (Fig. 16 in the appendix). Fewer than 0.3% of matched malaria records were missing dates and also removed from the analysis.

**Figure 16.**
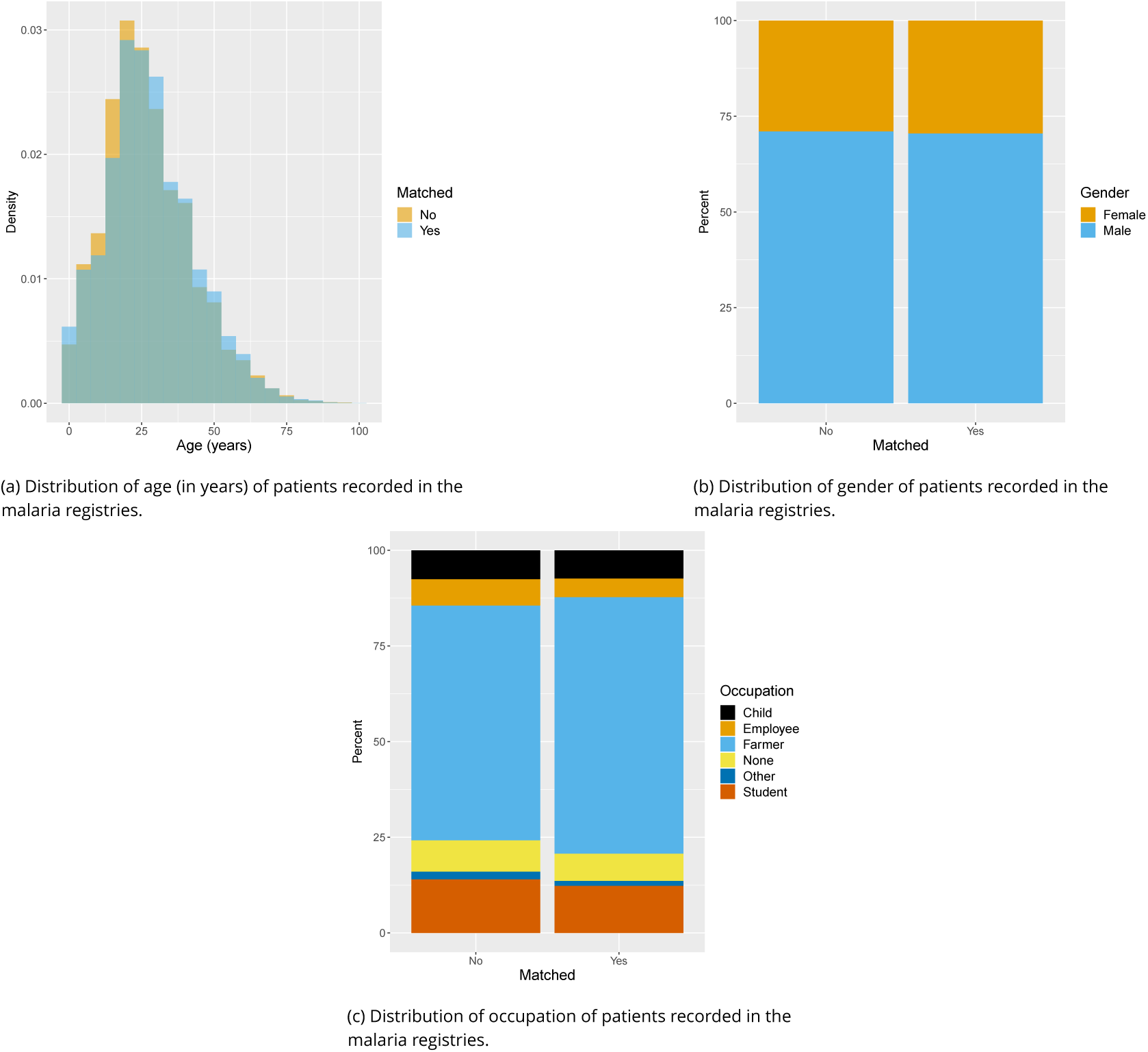
Additional figures from malaria registries: matched vs unmatched SES variables

### Spatio-temporal analysis

#### Deforestation

Table 1 and Figure 3 show the incidence rate ratio (IRR) associated with deforestation, measured by a 0.1 % increase in the percent area that experienced forest loss, in the previous 1 to 5 years within 1,10 and 30 km of villages.

Deforestation within 1 or 10 km of a village was not associated with malaria incidence rate in either the south or the north, regardless of the temporal lag. However, deforestation within 30 km of a village in the previous 1 and 2 years was associated with higher malaria incidence rates (e.g. 1-year lag, IRR = 1.23, 95% CI: [1.16; 1.3] in the south; IRR = 1.02, 95% CI: [0.94; 1.1] in the north) whereas deforestation in the previous 3,4 or 5 years was associated with approximately a 5% lower malaria incidence rate (e.g. 4-year lag, IRR = 0.94, 95% CI: [0.91; 0.97] in the south; IRR = 0.96, 95% CI: [0.92; 0.99] in the north).

These results suggest deforestation around villages, but not in the near vicinity (1 or 10 km), is associated with higher risk of malaria in the first two years and lower risk of malaria beyond. There was stronger evidence of associations with deforestation in the south, where IRRs were higher than in the north.

The IRR effect estimates in Table 1 and Figure 3 assume a linear relationship between deforestation and malaria. Figure 10 in the appendix shows a few of these relationships when such linearity isn’t assumed in the models. The functional forms reveal that they can be reasonably well summarized linearly, especially in the south. In the north, the functional forms highlight potential non-linearities for long term temporal lags but come with wide confidence intervals at low levels of deforestation. When statistically testing for non-linearities, the ANOVA *χ*^2^ test for goodness-of-fit was significant (5 % level) for only one of the 15 possible spatio-temporal scales specifications (30-km radius and 3-year temporal lag).

**Table 1.**
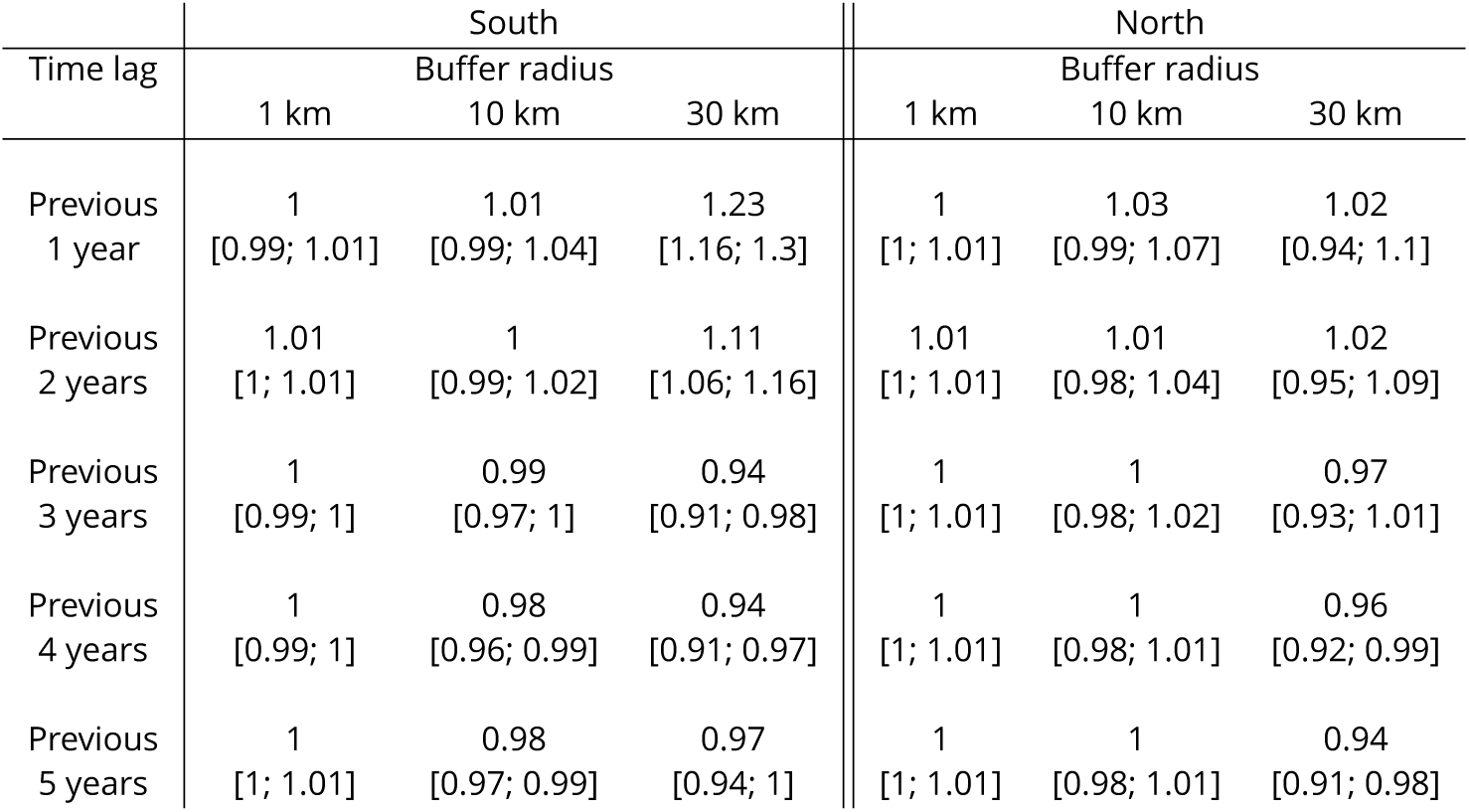
IRR between malaria incidence and a 0.1% increase in the area that experienced deforestation within 1, 10 or 30 km (left-right) of a village in the previous 1 to 5 years (top-down) in northern and southern Lao PDR. Adjusted for the spatio-temporal structure of the data, the environmental covariates selected in the model and forest cover within 30 km in the year before the deforestation temporal scale considered.

| Time lag | South |  |  | North |  |  |
| --- | --- | --- | --- | --- | --- | --- |
|  | 1 km | Buffer radius<br>10 km | 30 km | 1 km | Buffer radius<br>10 km | 30 km |
| Previous 1 year | 1<br>[0.99; 1.01] | 1.01<br>[0.99; 1.04] | 1.23<br>[1.16; 1.3] | 1<br>[1; 1.01] | 1.03<br>[0.99; 1.07] | 1.02<br>[0.94; 1.1] |
| Previous 2 years | 1.01<br>[1; 1.01] | 1<br>[0.99; 1.02] | 1.11<br>[1.06; 1.16] | 1.01<br>[1; 1.01] | 1.01<br>[0.98; 1.04] | 1.02<br>[0.95; 1.09] |
| Previous 3 years | 1<br>[0.99; 1] | 0.99<br>[0.97; 1] | 0.94<br>[0.91; 0.98] | 1<br>[1; 1.01] | 1<br>[0.98; 1.02] | 0.97<br>[0.93; 1.01] |
| Previous 4 years | 1<br>[0.99; 1] | 0.98<br>[0.96; 0.99] | 0.94<br>[0.91; 0.97] | 1<br>[1; 1.01] | 1<br>[0.98; 1.01] | 0.96<br>[0.92; 0.99] |
| Previous 5 years | 1<br>[1; 1.01] | 0.98<br>[0.97; 0.99] | 0.97<br>[0.94; 1] | 1<br>[1; 1.01] | 1<br>[0.98; 1.01] | 0.94<br>[0.91; 0.98] |

#### P. falciparum and P. vivax

One of the main differences between northern and southern Lao PDR with respect to our study is the malaria species composition. In the north, *P. vivax* is more prevalent with only a few sporadic and seasonal *P. falciparum* infections, whereas *P. falciparum* and *P. vivax* are co-endemic in the south (Fig. 2). We used this co-endemicity and the larger amount of malaria case data collected in the south to assess the relationship between deforestation and malaria for both species.

Table 2 and Figure 4 show that the pattern of spatio-temporal associations identified in Table 1 is primarily driven by *P. falciparum*, with no associations for deforestation in the near vicinity of villages (1 or 10 km) but a short-term increase (e.g. 1-year lag, IRR = 1.36, 95% CI: [1.25; 1.47]) and long-term decrease (e.g. 4-year lag, IRR = 0.86, 95% CI: [0.82; 0.9]) in *P. falciparum* malaria incidence for deforestation within 30 km of villages.

On the other hand, all the associations were attenuated for *P. vivax* infections. In the previous 2 years and within 30 km of villages, deforestation is still associated with a higher incidence of *P. vivax* (e.g. 1-year lag, IRR = 1.16, 95% CI: [1.09; 1.23]) but less so than for *P. falciparum*. However, regardless of the temporal lag or spatial scale, deforestation was no longer associated with lower *P. vivax* malaria risks.

Figure 22 in the appendix plots the species-specific relationships when not assuming linearity in the models.

**Table 2.**
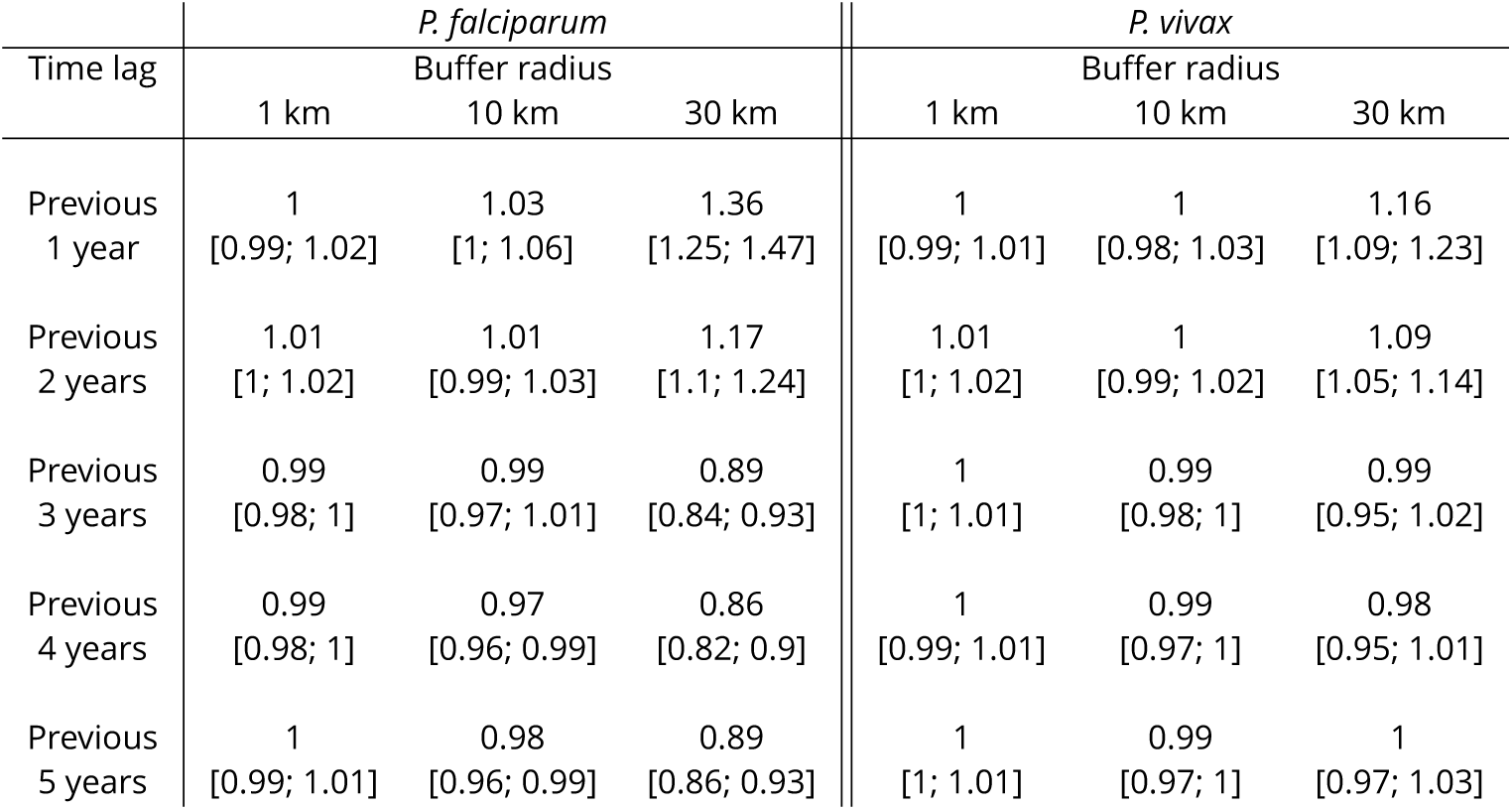
IRR between malaria incidence and a 0.1 % increase in the area that experienced deforestation within 1, 10 or 30 km (left-right) of a village in the previous 1 to 5 years (top-down) in southern Lao PDR, differentiated by malaria species. Adjusted for the spatio-temporal structure of the data, the environmental covariates selected in the model and forest cover within 30 km in the year before the deforestation temporal scale considered.

#### Alternative definitions of deforestation and interaction with forest cover

In previous models, our definition of deforestation did not distinguish between forest losses in densely forested areas and less forested areas. To explore potential interactions between deforestation and baseline forest cover, Table 3 shows how the IRR estimates vary as we consider deforestation in more densely forested pixels only (tree crown cover over 68% and 87% - see Methods for rationale on thresholds). We conducted this secondary analysis only for the non-null relationships previously identified, i.e. when considering a 30 km buffer radius around villages.

The associations with deforestation became more pronounced as we restricted forest losses to more forested areas: the IRR for deforestation in the previous 1 year, within 30 km of southern villages, increased from 1.23 (95% CI: [1.16; 1.3]) to 1.4 (95% CI: [1.05; 1.88]) when considering deforestation in areas with more than 0% and 87% tree crown cover respectively. On the other hand, the IRR for deforestation in the previous 4 years, within 30 km of southern villages, decreased from 0.94 (95% CI: [0.91; 0.97]) to 0.86 (95% CI: [0.79; 0.94]) when considering deforestation in areas with more than 0% and 87% tree crown cover respectively. A similar trend was observed in the north, although statistical significance wasn’t reached as frequently as in the south.

These results suggest that deforestation in deep and dense forests is more closely associated with malaria incidence in villages than deforestation in less forested areas.

**Table 3.** IRR between malaria incidence and a 0.1 % increase in the area that experienced deforestation within 30 km of a village in the previous 1 to 5 years (top-down) and within areas with tree crown cover density above 0%, 68% and 87% (left-right) in Lao PDR. Adjusted for the spatio-temporal structure of the data, the environmental covariates selected in the model and forest cover within 30 km in the year before the deforestation temporal scale considered.

| Time lag | South |  |  | North |  |  |
| --- | --- | --- | --- | --- | --- | --- |
|  | Deforestation within areas with tree crown cover density above |  |  | Deforestation within areas with tree crown cover density above |  |  |
|  | 0% | 68% | 87% | 0% | 68% | 87% |
| Previous 1 year | 1.23<br>[1.16; 1.3] | 1.45<br>[1.22; 1.72] | 1.4<br>[1.05; 1.88] | 1.02<br>[0.94; 1.1] | 1.08<br>[0.96; 1.2] | 1.36<br>[0.92; 2] |
| Previous 2 years | 1.11<br>[1.06; 1.16] | 1.13<br>[1.05; 1.22] | 1.33<br>[1.13; 1.56] | 1.02<br>[0.95; 1.09] | 1.02<br>[0.94; 1.1] | 1.02<br>[0.79; 1.31] |
| Previous 3 years | 0.94<br>[0.91; 0.98] | 0.92<br>[0.88; 0.96] | 0.88<br>[0.8; 0.97] | 0.97<br>[0.93; 1.01] | 0.96<br>[0.92; 1.01] | 0.87<br>[0.73; 1.02] |
| Previous 4 years | 0.94<br>[0.91; 0.97] | 0.93<br>[0.9; 0.96] | 0.86<br>[0.79; 0.94] | 0.96<br>[0.92; 0.99] | 0.95<br>[0.92; 0.99] | 0.84<br>[0.73; 0.96] |
| Previous 5 years | 0.97<br>[0.94; 1] | 0.95<br>[0.92; 0.99] | 0.92<br>[0.85; 1] | 0.94<br>[0.91; 0.98] | 0.94<br>[0.9; 0.98] | 0.79<br>[0.69; 0.9] |

## Discussion

Based on a large dataset of health facility surveillance records in two regions of Lao PDR, we found evidence that deforestation around villages is associated with higher malaria incidence over the short-term but lower incidence over the long-term (e.g, in the south, within 30 km of villages: IRR = 1.23 [1.16; 1.30] for deforestation in the previous year and IRR = 0.94 [0.91; 0.98] for deforestation in the previous 3 years). Our evaluation of alternative spatial scales identified strong associations for deforestation within a 30 km radius around villages but not for deforestation in the near (10 km) and immediate (1 km) vicinity. Our results incorporated correction for the probability of seeking treatment, modeled as a function of distance to the closest health facility, as well as adjustment for several environmental covariates. Results appear driven by deforestation in densely forested areas and the patterns exhibited are clearer for infections with *P. falciparum* than for *P. vivax*.

The wide availability and longitudinal nature of malaria surveillance records collected routinely by the national program enabled exploration of the relationship between deforestation and malaria incidence over multiple spatio-temporal scales and across different levels of forest density. The spatio-temporal variability highlighted here provides insights into the causal mechanisms driving local-scale malaria incidence in the GMS. This approach not only quantified the deforestation-malaria incidence association in the GMS, but also strengthened the evidence for the key influence of forest-going populations on malaria transmission in the GMS.

Results from this study echo the frontier malaria hypothesis originating from the Amazon region, which posits an increase in malaria incidence in the first few years following deforestation and a decrease over the long term. In our study, however, the time scale for the inflexion point was shorter, 2-3 years after deforestation compared to 6-8 years in the Amazon [16], most likely because of very different underlying human processes. Indeed, the frontier malaria hypothesis considers non-indigenous human settlements sprouting deeper and deeper in the forest whereas forest-going populations in the GMS are primarily members of established forest-fringe communities who regularly tour the forest overnight to hunt and collect wood [21]. Industrial and agricultural projects or lucrative forest-based activities also attract mobile and migrant populations (MMPs) [30] in remote forested areas of the GMS but not on the same scale as the politically and economically driven unique colonization of the Amazon [16].

Our results are also consistent with the three previous multivariable empirical studies [27, 53, 25] that assessed the link between deforestation and malaria in Southeast Asia. Our study builds on these findings by using higher resolution forest data and exploring additional spatio-temporal scales. Using biennial village census data from Indonesia between 2003 and 2008 and district-aggregated remote sensing forest data, Garg [27] reported a 2 to 10.4% increase in the probability of a malaria outbreak in each village of districts that lost 1000 hectares of their forest cover in the same year. Using data from a 1996 cross-sectional household survey conducted in a quasi-experimental setting around a protected area in Indonesia, Pattanayak et al. [53] found a positive association between disturbed forest (vs undisturbed) and malaria in children under 5, again using no temporal lag. Our analysis plan was largely inspired by Fornace et al. [25], which used similar high-resolution forest data [34] and 2008-2012 incidence data from Sabah, Malaysia. They reported a 2.22 (95% CI: [1.53; 2.93]) increase in the *P. knowlesi* incidence rate for villages where more than 14% (< 8%, being the reference) of the surrounding area (within 2 km) experienced forest loss in the previous 5 years. On the other hand, our analysis explored wider spatial scales, bypassed any coarse categorization of forest and deforestation variables, corrected incidence for treatment-seeking probability, and most importantly focused on *P. falciparum* and *P. vivax*, the dominant malaria parasites in the GMS.

Engaging in forest activities, such as logging, hunting or spending the night in the forest, has been reported as a major risk factor by many studies in the region [17, 20, 39, 22, 61]. As countries of the GMS work towards malaria elimination, the literature stresses the key role of forest-going populations [30, 45,11, 68, 56], although research programs highlight the challenges of accessing them [14, 40] as well as their diversity [45,11]. To our knowledge, no previous study has leveraged geo-spatial statistical analyses to characterize the importance of forest-going populations in the GMS. Our results suggest that deforestation in dense forests (Table 3) around villages, particularly areas further from the village (Table 1), is a driver of malaria in Lao PDR. We argue that this is indicative of the existence of a key high-risk group linking the deforestation patterns identified to malaria in the villages, namely a forest-going population. Deforestation captured by remote sensing in this setting likely reflects locations and times of heightened activity in the forest areas near villages, and therefore greater human-vector contact. We suspect longer and deeper trips into the forest result in increased exposure to mosquitoes, putting forest-goers at higher risk.

We conducted this study in northern and southern Lao PDR, where the malaria species composition differs, and assessed species-specific relationships in the south where *P. falciparum* and *P. vivax* are co-endemic. Our results highlight the challenges ahead of national programs with *P. vivax* elimination after successful *P. falciparum* elimination, as increasingly mentioned in the literature [18, 38]. This study identified a clear pattern of spatio-temporal associations between *P. falciparum* and deforestation, but these were not apparent for *P*. *vivax* (Table 2). The increase in *P*. *vivax* incidence in the first 2 years following deforestation was identified as well but the associations were smaller than for *P. falciparum*. Importantly, deforestation was never associated with lower risks of *P*. *vivax*. A recent study in the Amazon [43] reported a similar attenuation of the effects of deforestation on *P*. *vivax* compared to P *falciparum*, most likely because of *P*. *vivax* parasites’ ability to relapse months or even years after infection, which decouples the association between transmission and incidence data. These species-specific differences may also explain why the pattern of spatio-temporal associations between malaria and deforestation were markedly clearer in the south than in the north where *P*. *vivax* dominates.

Our results did have some inherent limitations based upon routine health facility surveillance data. First, completeness and reliability of health facility surveillance records is highly variable across and within countries of the GMS and may depend on malaria incidence level, as resources tend to be prioritized on high-prevalence diseases. Second, another challenge with these data is obtaining an accurate denominator for incidence, as not everyone attends a public health center when febrile. We addressed this issue by modeling the probability of seeking treatment as a function of travel time to the closest health facility using data from two cross-sectional surveys. Third, the village-level geo-referencing of malaria registries ignores the possibility that patients may become infected elsewhere. Unfortunately, these surveillance records did not include information about patients’ recent travels or forest trips. Research to track micro-scale movements of forest-goers is needed to understand how they interact with the forest and where are the foci of infection.

In addition, the forest data used, although it offers reproducible, well-defined and high-resolution forest variables, has also been criticized, in particular for not distinguishing tropical forests from agroforestry [68, 36]. Finally, our relative measure of deforestation, key to consistently compare the effects across different spatial scales, also implies that a 0.1 % of the area that experienced forest loss within 30 km of a village is a much larger area (~ 280 hectares) than within 1 km (~ 0.3 hectare) and should be interpreted cautiously.

In conclusion, this study assessed the relationship between deforestation and malaria in Lao PDR. Our approach leveraged surveillance records collected by the national malaria program and high-resolution forest data and rigorously explored the spatio-temporal pattern of associations. As countries of the GMS work towards malaria elimination, our results highlight the challenges to transition from *P. falciparum* to *P*. *vivax* elimination, confirm and characterize the importance of high-risk populations engaging in forest activities and suggest malaria programs may benefit from monitoring areas of on-going deforestation using remotely sensed data.

## Materials and methods

### Study site and population

This study was conducted in eight districts (Fig. 5) to leverage the ecological and epidemiological diversity of Lao PDR. Four districts (Moonlapamok, Pathoomphone, Sanasomboon and Sukhuma) are situated in the southern province of Champasak where both *P*. *falciparum* and *P. vivax* are endemic. The four other districts (Et, Paktha, Nambak and Khua) each come from one of four northern provinces (Bokeo, Huaphanh, Phongsaly, Luang-Prabhang) where *P*. *vivax* is endemic but *P*. *falciparum* has reached historical lows [2].

The four districts in the north were chosen in consultation with district and provincial level malaria staff to represent the epidemiology of malaria in northern Lao PDR. They were selected as part of a cross-sectional survey designed to assess the prevalence and risk factors for malaria in northern Lao PDR [41]. This region is very mountainous and characterized by a diverse climate, low-population density and limited road access [7].

The 4 districts in the south were selected within a larger cluster randomized controlled trial (RCT) study designed to assess the effectiveness of high-risk group targeted active case detection in southern Lao PDR [40], where more than 95% of the country malaria burden is concentrated [2]. This region is characterized by a moderately hilly and forested terrain and a workforce primarily engaged in forest-based and agricultural activities [14].

### Malaria data

#### Malaria case data

We conducted a retrospective review of malaria registries recorded at all health centers in the study districts between January 2013 and December 2016 in the north and between October 2013 and October 2016 in the south. The registries included information on every patient that was tested (RDT and/or microscopy) for malaria at the health center. Date, species-specific test results, demographic variables (age, gender and occupation) and the village of residence of the patient were recorded in the registries. With help from local Lao experts, village names were matched to a geo-registry of all villages in Lao PDR compiled from the 2005 and 2015 national census [1] and provided by the Center for Malariology, Parasitology and Entomology (CMPE). The geo-registry contains GPS coordinates and population of Lao PDR’s villages. Unmatched records and records with missing date were removed from the analysis. Finally, these data were aggregated to extract the monthly village-level malaria incidence.

#### Treatment-seeking data

One issue with using passive surveillance data is that not everyone will seek treatment at a public health facility for a febrile illness, which can lead to an underestimate of the true incidence, if not accounted for. To correct for that, we modeled the probability that an individual in a given village of the study’s district would seek treatment at a public health facility when febrile. We assumed that such probability is essentially driven by the travel time to the closest health facility. See supplementary material S1 for methods used to calculate travel times to closest health facilities.

To model the probability of seeking treatment, we used data from two cross-sectional household surveys conducted in the eight districts where registries were collected. In the north, 1,480 households across 100 villages were surveyed in September-October 2016 [41]. In the south, 1,230 households across 56 villages were surveyed in the baseline assessment of the RCT [40] in December 2017. In particular, survey respondents were asked whether or not they would seek treatment at the closest health facility for a febrile illness and GPS coordinates of their household were recorded.

We then used the cross sectional surveys to model the probability of seeking treatment (at a public health facility, implicit from now on), *θ*, as a function of travel time to the closest health facility, *τ* (Equation 1). To account for the correlation structure induced by the stratified sampling approach used in the surveys, we modeled the number of successes (febrile patients seeking treatment), *S_hv_*, at the household level and included a random intercept for village in the logistic regression.

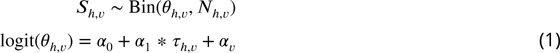

where *N_h,v_* is the number of febrile individuals in household h of village v and 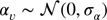.

We fit the models separately in the north and in the south and used the region-specific model to predict the probability of seeking treatment at all villages of the study districts based on their distance to the closest health facility. The population who seek treatment was then calculated by multiplying the village population by the probability of seeking treatment. See supplementary material S2 for travel times and treatment-seeking probabilities results.

### Forest data

For every 30m pixel in Lao PDR, tree crown cover density for the year 2000 and year of forest loss between 2000 and 2017, were obtained from Hansen et al. [34] (See illustrative example in pixels 1 and 2 of Fig. 6). These layers were produced using decision tree classifiers on Landsat remote sensing imagery [34]. Trees are defined as “all vegetation taller than 5m in height” and forest loss as “stand-replacement disturbance or the complete removal of tree cover canopy” [34], meaning “the removal or mortality of all tree cover in a Landsat pixel” [33].

#### Deforestation variable

To define our primary exposure variable, for all villages in the study districts and year of the study period, we calculated the percent area within a buffer radius of 1,10 and 30 km that experienced forest loss in the previous 1, 2, 3, 4 and 5 years (Fig. 6). These distances were chosen to explore a range of spatial scales at which the forest environment may be differentially relevant for village-based populations and forest-goers. To explore potential interactions between deforestation and forest cover, we computed an alternate exposure variable, restricting to areas that both experienced forest loss and had a tree crown cover density above 68 and 87%. Those thresholds are limits of the inter-quartile range (IQR) of the distribution of tree crown cover density in any 30m pixels within 10 km of study’s villages that experienced forest loss between 2000 and 2017. They represent deforestation activities occurring in areas of increasing forest cover.

#### Forest cover variable

We also combined the two Hansen layers to produce annual tree crown cover maps of the study districts, assuming no changes prior to the year of forest loss but setting to 0 the pixel tree crown cover density afterwards (Fig. 6). For all villages in the study districts and year of the study period, we calculated the average tree crown cover density within a buffer radius of 1,10 and 30 km and for 0,1, 2 and 3-year lags. This is a secondary exposure and will be adjusted for in the primary analysis.

### Environmental covariates

Village population sizes were needed to estimate monthly malaria incidence. 2005 and 2015 population estimates for the 491 villages of study districts were obtained from the national census [1]. The annual population growth rate (3.7%) was used to impute population values for two villages missing 2005 estimates and for two villages missing 2015 estimates. Then, village-level population growth rates were used to estimate villages’ population per year between 2008-2016, assuming linear annual growth rate (median = 1.7%, IQR = [0%; 4.5%]).

Altitude, temperature, rainfall and access to health care were considered as potential village-level confounders of the relationship between malaria and forest cover factors. Travel time to closest health facility, computed for the treatment-seeking model, was used as a proxy for health care access. Altitude was extracted from SRTM [37] 1 km resolution layers. Monthly average day and night temperature were extracted from MODIS 1 km resolution product (MOD11C3 [66]). Finally, monthly total rainfall was extracted from CHIRPS [26] 1 km resolution publicly available data. For every month of the study period, monthly temperatures and precipitation in the previous 1, 2 and 3 months were also extracted as well as the average temperatures and total precipitation over the previous 1, 2 and 3 months. Annual total precipitation and its seasonality (coefficient of variation), as well as the annual mean temperature and its seasonality (standard deviation), were also extracted from the WorldClim [24] 1 km resolution datasets.

Altitude was missing for one village and we used an online elevation finder tool (FreeMapTools) for imputation. Temperature was missing for 2.4% of the village-months over the study period, most likely because of cloud coverage of the MODIS imagery. Monthly temperature was never missing more than two years in a row at villages of the study’s districts and we imputed the temperature of the same month of the following year (or prior year when needed), adjusting for average district-level monthly temperature differences between the two consecutive years. Monthly rainfall and WorldClim data were not missing at any of the villages.

### Statistical analysis

#### Statistical model

To model malaria incidence (Equation 2), the number of positive cases *Y_v,t_* at village v over month t was modeled using a generalized additive model (GAM) [69]. To account for overdispersion, a negative binomial distribution was used, including an additional variance parameter *v*. The probability of seeking treatment *θ_v_*, estimated from the treatment-seeking model, was multiplied by the village population *Pop_v,t_* to derive the population seeking treatment,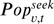. This was included as an offset term in the incidence model. Spatial autocorrelation was accounted for by the bivariate thin plate spline smoothing function on coordinates, *f* (*Lat, Long*) and village random intercepts were included. A non-linear temporal trend was also included with the smoothing function on month, *f*(*t*). Finally, the primary exposure, deforestation, and potential environmental confounders, including forest cover, were modeled with splines in 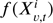for maximum flexibility. See Figure 8 in supplementary material S3 for a graphical visualization of our conceptual model for this analysis.

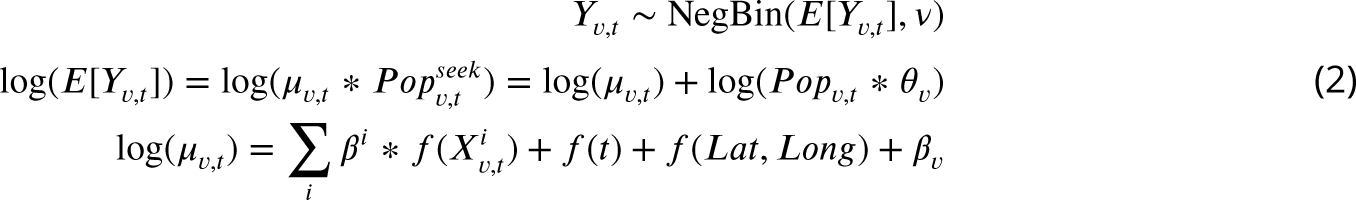

with 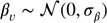.

After model selection of the best fitting combination of covariates, 15 models were run separately in the north and the south, each varying the buffer radius (1,10 and 30 km) and temporal scale for deforestation (previous 1, 2, 3, 4 and 5 years). The coefficients of the linear effect for deforestation were extracted and exponentiated to get the incidence rate ratio (IRR) associated with a 0.1% increase in the percent area that experienced forest loss around villages.

As secondary analyses, we ran the same models, separately by malaria species *(P. falciparum* and *P. vivax)*. Finally, we used our alternative definitions for deforestation, restricted to areas that both experienced forest loss and had a tree crown cover density above 68 and 87%, to explore the interaction between deforestation and the amount of forest cover.

#### Model selection

With no pre-specification for the spatio-temporal scale at which the effect of deforestation might affect malaria incidence, it was important to be able to compare the effect estimates across models with varying deforestation variables. We therefore looked for the combination of potential covariates that provided the best fit and only then included the varying deforestation variables, hence assessing the adjusted effect estimate of deforestation consistently across models. The entire model selection process was carried using data from the south only and modeling non-species specific malaria incidence. This larger dataset provided the most statistical power to fit flexible relationships and identify a core way to adjust for covariates.

First, for each of the three monthly environmental variables (precipitation, day and night temperature), we selected the one of its seven variations (in current month, in previous 1, 2 or 3 months and aggregated over current and previous 1, 2 or 3 months) that provided the best AIC fit in an univariate model, solely adjusted for the spatio-temporal structure of the data *(f*(*t*), *f*(*Lat, Long*) and village random intercepts).

Second, we used a modified version of the forward and backward selection approaches to build the best AIC fitting combination of the nine potential covariates (three monthly variables selected, four WorldClim variables, altitude and travel time to nearest health facility), still adjusting for the spatio-temporal structure of the data. As in the classical selection approaches, variables were added and dropped one at a time, but were retained based on the largest improvement (decrease) in AIC and not p-value dependent. The model obtained by the forward selection approach had the lowest AIC and was therefore retained. Average night temperature over the current month and previous 3 months, average day temperature over current month, total precipitation in current month, total annual precipitation and its seasonality (coefficient of variation) and travel time to the closest health facility made it into the selected model.

Third, we ran 12 models, each varying the buffer radius (1,10 and 30 km) and temporal scale (0,1, 2 and 3 year lag) for the forest cover variable, on top of the previously selected model. The coefficients of the linear effect for forest cover were extracted and exponentiated to get the incidence rate ratio (IRR) associated with a 1 % increase in the average tree crown cover density around villages (Table 7 in the appendix). The model including average tree crown cover density within 30 km of villages with no temporal lag provided the smallest AIC value.

In the final models with the deforestation variables we therefore included the average tree crown cover density within 30 km of villages in the starting year of the temporal scale for the deforestation variable considered (e.g. 3 year lag in the model with percent area that experienced forest loss in previous 3 years as the deforestation variable) to adjust for baseline forest cover.

#### Sensitivity analyses

Two sensitivity analyses strengthening the robustness of our primary analysis were conducted. In the first one, villages’ populations at risk of appearing in the surveillance system registries were not adjusted for the probability of seeking treatment. In the second one, we further adjusted our final models for the village malaria incidence rate in the previous month. See supplementary materials S4.

## Ethics

This study was approved by the National Ethics Committee for Health Research at the Lao Ministry of Health (Approval #2016-014) and by the UCSF ethical review board (Approvals #16-19649 and #17-22577). The informed consent process was consistent with local norms, and all study areas had a consultation meeting with, and approvals from, village elders. All participants provided informed written consent; caregivers provided consent for all children under 18, and all children aged 10 and above also provided consent directly. The study was conducted according to the ethical principles of the Declaration of Helsinki of October 2002.

## Data Availability

All data generated and analyzed in this study have been uploaded with the submission.

## Acknowledgments

For their expertise and assistance, we thank Michelle Roh (Study design and interpretation), Ricardo Andrade Pacheco (GAM and spatial modeling), Alemayehu Midekisa (Remote sensing), Stephen Shiboski (Statistical analyses) and Maria Glymour (Causal inference and manuscript writing). FRwas funded was the Bill & Melinda Gates Foundation (Grant ID OPP1116450).

## Supplementary Materials

### S1: Travel times methods

To calculate the travel time along a path linking any two points of the map, we defined a transition matrix that gives the speed at which one may travel between two adjacents pixels. We followed the parameterization suggested by Alegana et al. [9] and demonstrated by Sturrock et al. [58] (see Table 4), which first uses Toblers’ hiking function to specify the travel speed between two points of different altitudes. Intuitively, it is faster to travel downhill than uphill. Second, the speed is adjusted based on the type of landcover travelled through: a forested or a flooded area for instance slows you down. Last, the network of roads and major rivers may be used to catch a bus or a boat and therefore increases the travel speed. Altitude (SRTM 90m [37]) was aggregated and resampled at the land cover (ESA GlobCover 2009 Project [5]) 300m-resolution and roads and waterways from Open Street Map [3] were rasterized to calculate the transition matrix all across Lao PDR. The ‘raster’ package in R [4] was used.

**Table 4.** Data used to parameterized the transition matrix with the travel speed between any 2 adjacent pixels of the map

| Data layer | Category | Speed (km/h) |
| --- | --- | --- |
| Digital elevation (slope) | 0°(flat) | 5 |
|  | 5°(uphill) | 3.71 |
|  | -5°(downhill) | 5.27 |
| Land cover | Cropland | No adjustment |
|  | Artificial and bare areas | No adjustment |
|  | Open deciduous forest | 0.8 * Hiking speed |
|  | Sparse herbaceous | 0.8 * Hiking speed |
|  | Closed deciduous forest | 0.6 * Hiking speed |
|  | Herbaceous | 0.6 * Hiking speed |
|  | Flooded | 0.5 * Hiking speed |
|  | Other forest cover | 0.4 * Hiking speed |
|  | Water | 0.2 * Hiking speed |
| Roads and rivers | Motorway/trunk | 80 |
|  | Primary/secondary | 60 |
|  | Tertiary/unclassified | 10 |
|  | Major rivers | 5 |

We then used the Djisktra’s algorithm from the R-package igraph [19] and the gdistance package [23] to find the fastest route between every village (or every household in the cross-sectional surveys) and its closest health facility. Coordinates of health facilities across Lao PDR came from the 2017 stratification exercise and were provided by CMPE. We authorized travel through non-study districts but not across international borders.

### S2: Travel times and treatment-seeking results

Figure 7a shows how the travel time to closest health facility varies across Champasak province in southern Lao PDR, influenced by both distance and road connectivity. Figure 7c presents a right-skewed distribution of travel time from study villages to the closest health facility. Most villages are within 2 hours of the closest health facility but some are as far as 6 hours away. The distribution is similar for villages in the northern and southern study districts.

In the southern household survey, 243 individuals reported fever in the past 2 weeks. 225 (92.6%) of them, from 156 households, answered whether or not they sought treatment and were included in the treatment-seeking model. 219 (97.3%) reported seeking treatment and they all reported where they did so: 154 (70.3%) of them sought treatment at a public health facility (Village malaria worker (VMW), health center, district hospital or provincial hospital) and would therefore appear in the malaria registries collected.

In the northern household survey, 378 individuals reported fever in the past 2 weeks. 360 (95.2%) of them, from 297 households, answered whether or not they sought treatment and were included in the treatment-seeking model. 283 (78.6%) reported seeking treatment. Only 40 (14.1%) of them reported where they did so but all of them sought treatment at a public health facility and we therefore upweighted the population that sought treatment at a public health facility accordingly.

Most surveyed households included in the treatment-seeking model were within 2 hours of travel time to the closest health facility but some were almost 5 hours away (Fig. 17 in the appendix). Figure 7b shows the modeled relationship between the probability of seeking treatment (at a public health facility, implied from now on) and distance to the closest health facility. For villages within the same 300m^2^ pixel as a health facility (estimated travel time of 0 hour), the predicted probability of seeking treatment is 0.87 (95% CI: [0.79; 0.92]) in the north and 0.78 (95% CI: [0.63; 0.89]) in the south. A 1 hour increase in travel time to the closest health facility is associated with a similar 0.79 (95% CI: [0.55; 1.13]) reduction in the odds of seeking treatment in the north and 0.76 (95% CI: [0.43; 1.34]) in the south, almost reaching statistical significance when pooling data from both regions: 0.77 (95% CI: [0.56; 1.04]). Figure 7d shows the resulting distribution for the probability of seeking treatment for all villages in study’s districts. Monthly village-level malaria incidence was adjusted accordingly.

**Figure 17.**
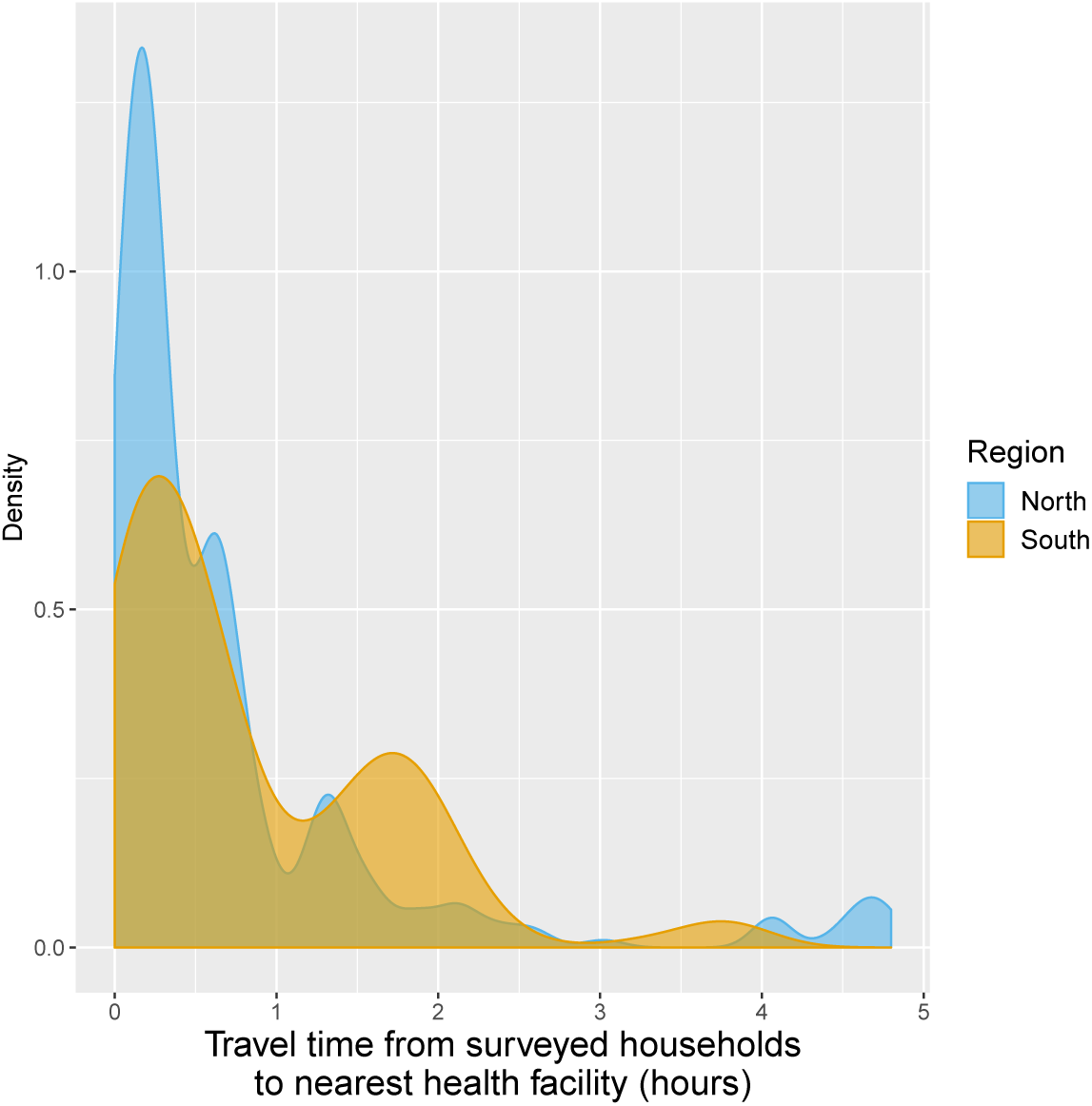
Distribution oftravel time (in hours) from surveyed households to closest health facilities.

### S3: Conceptual Model

### S4: Sensitivity Analysis

We conducted a first sensitivity analysis where village population at risk of appearing in the surveillance system registries were not adjusted for the probability of seeking treatment. The effect estimates and confidence intervals were virtually unchanged (Table 5 in the appendix).

Our second sensitivity analysis involved adjustment for the village malaria incidence rate in the previous month in our models. The effect estimates and confidence intervals were fairly unchanged, albeit a small reduction in the estimated short-term positive association between recent deforestation and malaria incidence (Table 6 in the appendix).

Overall, those sensitivity analyses strengthen the robustness of our primary analysis.

**Table 5.**
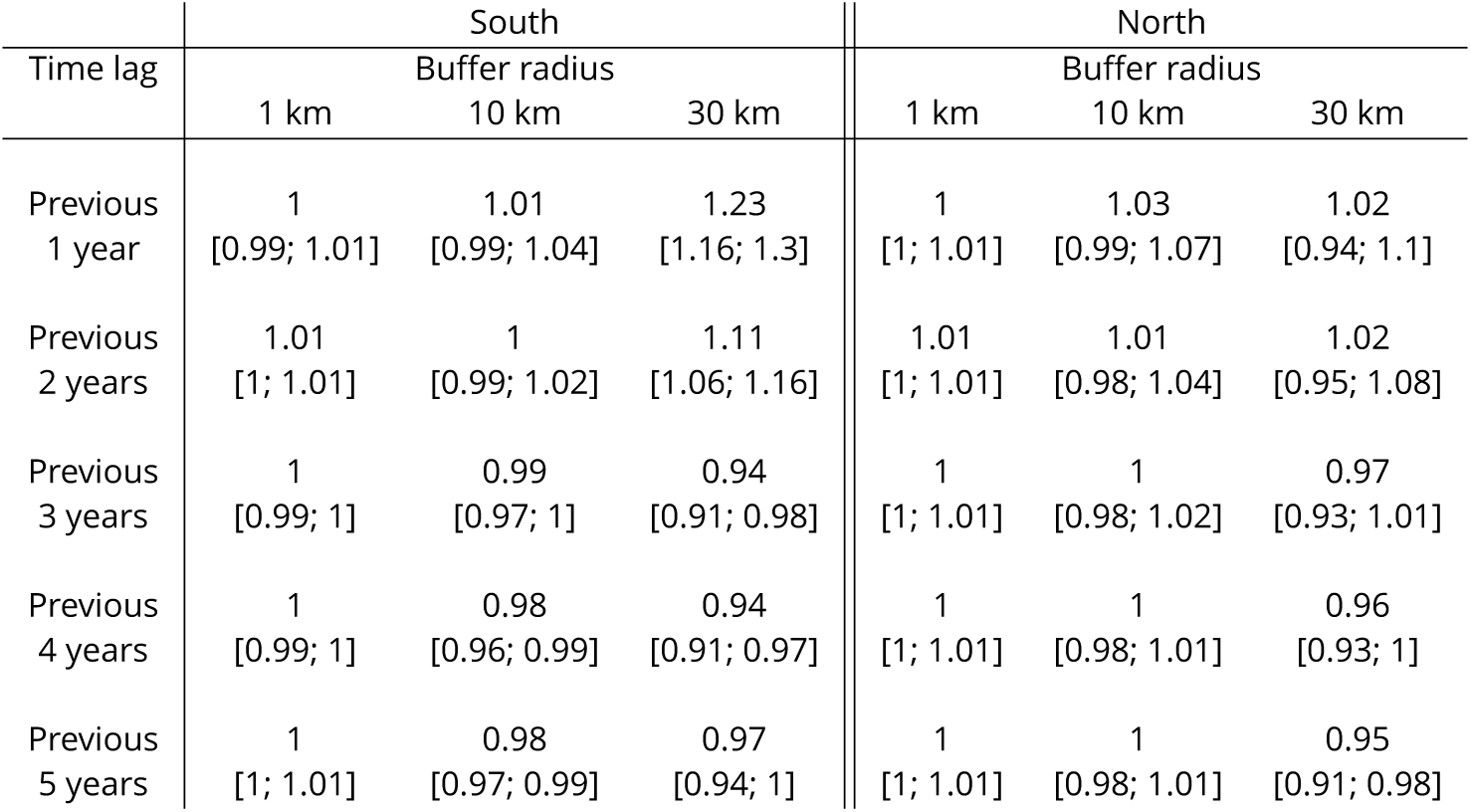
IRR associated with a 0.1% increase in forest loss. Adjusted for the spatio-temporal structure of the data, the environmental covariates selected in the model and forest cover within 30 km in the year before the deforestation temporal scale considered. Sensitivity analysis 1: village population unadjusted for probability of seeking treatment.

**Table 6.** IRR associated with a 0.1% increase in forest loss. Adjusted for the spatio-temporal structure of the data, the environmental covariates selected in the model and forest cover within 30 km in the year before the deforestation temporal scale considered. Sensitivity analysis 2: models adjusted for village monthly malaria incidence rate in previous month.

| Time lag | South |  |  | North |  |  |
| --- | --- | --- | --- | --- | --- | --- |
|  | 1 km | Buffer radius<br>10 km | 30 km | 1 km | Buffer radius<br>10 km | 30 km |
| Previous<br>1 year | 1<br>[0.99; 1.01] | 1.01<br>[0.99; 1.03] | 1.17<br>[1.1; 1.24] | 1<br>[1; 1.01] | 1.02<br>[0.98; 1.06] | 1<br>[0.92; 1.08] |
| Previous<br>2 years | 1<br>[1; 1.01] | 1<br>[0.98; 1.01] | 1.08<br>[1.04; 1.13] | 1<br>[1; 1.01] | 1.01<br>[0.99; 1.04] | 1.01<br>[0.95; 1.08] |
| Previous<br>3 years | 1<br>[0.99; 1] | 0.98<br>[0.97; 1] | 0.93<br>[0.9; 0.97] | 1<br>[1; 1.01] | 1<br>[0.98; 1.02] | 0.97<br>[0.93; 1.01] |
| Previous<br>4 years | 1<br>[0.99; 1] | 0.98<br>[0.96; 0.99] | 0.94<br>[0.91; 0.97] | 1<br>[1; 1.01] | 1<br>[0.99; 1.02] | 0.96<br>[0.93; 0.99] |
| Previous<br>5 years | 1<br>[1; 1.01] | 0.98<br>[0.96; 0.99] | 0.96<br>[0.93; 0.99] | 1<br>[1; 1.01] | 1<br>[0.99; 1.02] | 0.95<br>[0.92; 0.98] |

### S5: Additional Results

#### Environmental covariates

Figure 9 shows the relationship - via their individual contribution *β* * *f* (*X*) in equation 2 - between malaria incidence and the environmental covariates in the multivariable model selected by the selection approach: time to nearest health facility, average night temperature (over the current and previous 3 months), average day temperature (over the current month), total monthly precipitation (over current month) and average total annual precipitation and its coefficient of variation. We see that precipitation variables have the largest effect on malaria incidence and that relationships differ slightly by region although the range covered by the environmental variables also differs by region. Note that 95% confidence intervals (see Fig. 18 in the appendix) have been hidden for better visualization. The larger amount of data in the south allowed the identification of more precise relationships than in the north.

**Figure 18.**
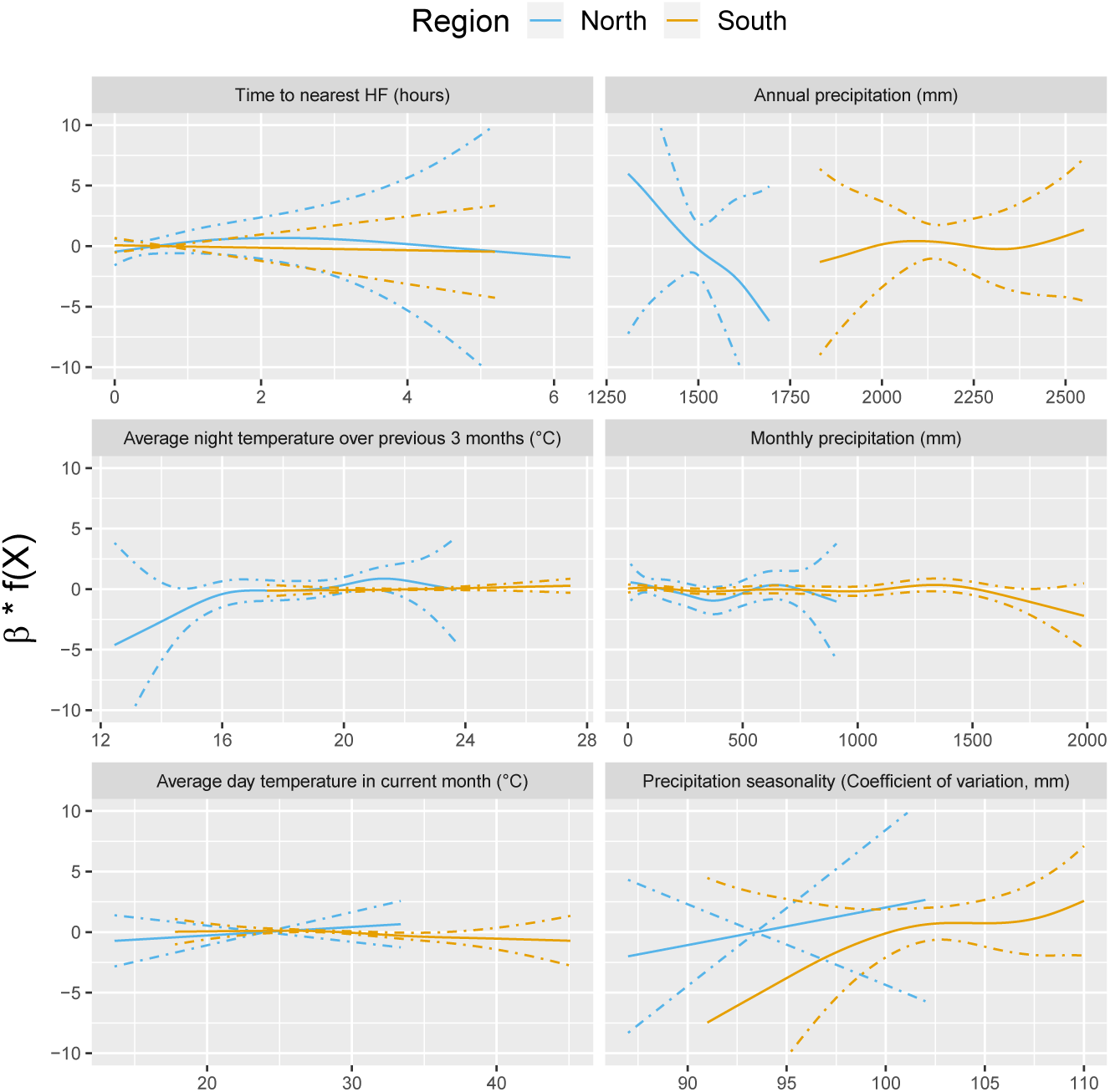
Relationships between malaria incidence and environmental covariates in the multivariable model selected by the forward selection approach, as described in the methods section. The model was additionally adjusted for the spatio-temporal structure of the data (*f*(*t*), *f* (*Lat, Long*) and village random intercepts). Dashed lines are for 95% confidence intervals. Note that the y scale has been trimmed for better visualization.

#### Forest cover

Table 7 shows the incidence rate ratio (IRR) associated with forest cover, measured by a 1 % increase in the average tree crown density, in current and previous 3 years within 1,10 and 30 km of villages.

Forest cover within 1 km of a village was not associated with malaria incidence rate in either the south or the north, regardless of the temporal lag. However within 10 and 30 km of a village, increased forest cover tended to be associated with higher malaria incidence rates both in the north and the south (e.g. 30 km buffer, 1-year lag, IRR = 1.1, 95% CI: [0.98; 1.24] in the south; IRR = 1.24, 95% CI: [1.05; 1.46] in the north). The associations were higher when considering a larger spatial scale (30 km) but were already statistically significant for a 10 km buffer radius in the south. The temporal scale considered did not affect the associations much.

Statistical significance wasn’t necessarily reached for all the associations highlighted but the trends observed suggest forest cover around villages but not in the immediate vicinity (1 km) leads to higher risk of malaria both in the north and in the south.

**Table 7.** IRR [95% CI] associated with a 1 % increase in average tree crown density. Adjusted for the spatio-temporal structure of the data and the environmental covariates selected in the model.

| Time lag | South |  |  | North |  |  |
| --- | --- | --- | --- | --- | --- | --- |
|  | 1 km | Buffer radius<br>10 km | 30 km | 1 km | Buffer radius<br>10 km | 30 km |
| Current year | 0.99<br>[0.98; 1.01] | 1.08<br>[1.05; 1.12] | 1.05<br>[0.94; 1.17] | 0.98<br>[0.95; 1.02] | 1.03<br>[0.96; 1.1] | 1.26<br>[1.05; 1.51] |
| Previous 1 year | 1<br>[0.98; 1.02] | 1.09<br>[1.05; 1.13] | 1.1<br>[0.98; 1.24] | 0.99<br>[0.96; 1.03] | 1.03<br>[0.96; 1.1] | 1.24<br>[1.05; 1.46] |
| Previous 2 years | 1<br>[0.98; 1.02] | 1.09<br>[1.05; 1.13] | 1.13<br>[1; 1.27] | 1<br>[0.97; 1.04] | 1.03<br>[0.96; 1.1] | 1.17<br>[0.98; 1.41] |
| Previous 3 years | 1<br>[0.98; 1.02] | 1.09<br>[1.05; 1.13] | 1.15<br>[1.02; 1.3] | 1.01<br>[0.97; 1.05] | 1.03<br>[0.96; 1.11] | 1.2<br>[1.02; 1.43] |

The model including average tree crown cover density within 30 km of villages with no temporal lag provided the best AIC. In the final models with the deforestation variables we therefore included the average tree crown cover density within 30 km of villages in the starting year of the temporal scale for the deforestation variable considered (e.g. 3 year lag in the model with percent area that experienced forest loss in previous 3 years as the deforestation variable).

#### Deforestation - Non linearities

The IRR effect estimates in Table 1 and Figure 3 assume a linear relationship between deforestation and malaria. Figure 10 shows a few of these relationships - via their individual contribution *β* * *f* (*X*) in equation 2 - when such linearity isn’t imposed in the GAM models. Although the AIC fit is slightly better when modeling non-linearities, we see that the linearity assumption is mostly warranted, especially in the south. A quadratic relationship may have been better suited in the north but the initial rise comes with a very large confidence interval, as a result of the lesser amount of data.

**Figure 19.**
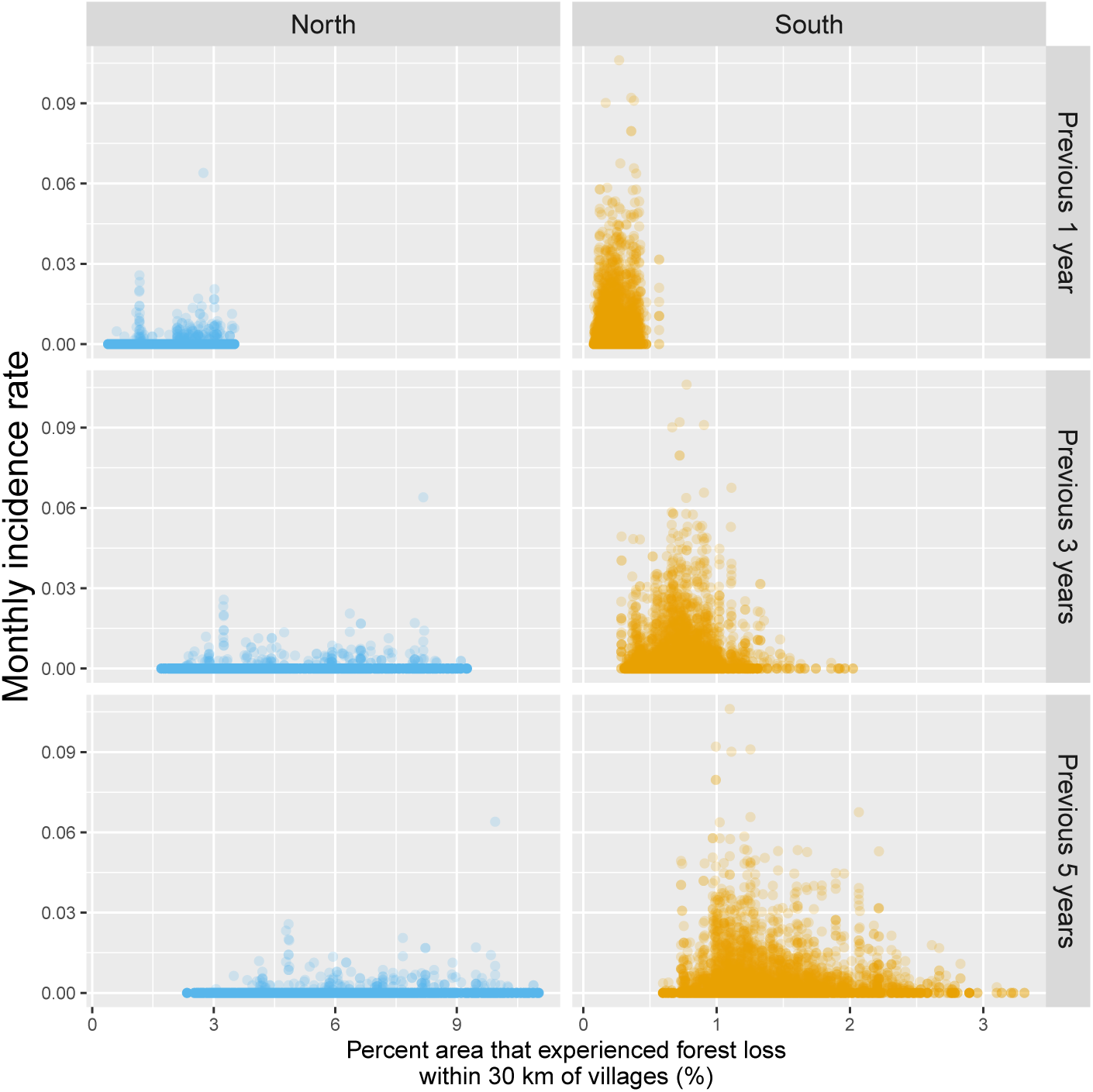
Raw scatterplot between monthly village malaria incidence rate and the percent area within 30 km of villages that experienced forest loss in the previous 1, 3 and 5 years. Note that scales are different between regions for better visualization.

**Figure 20.**
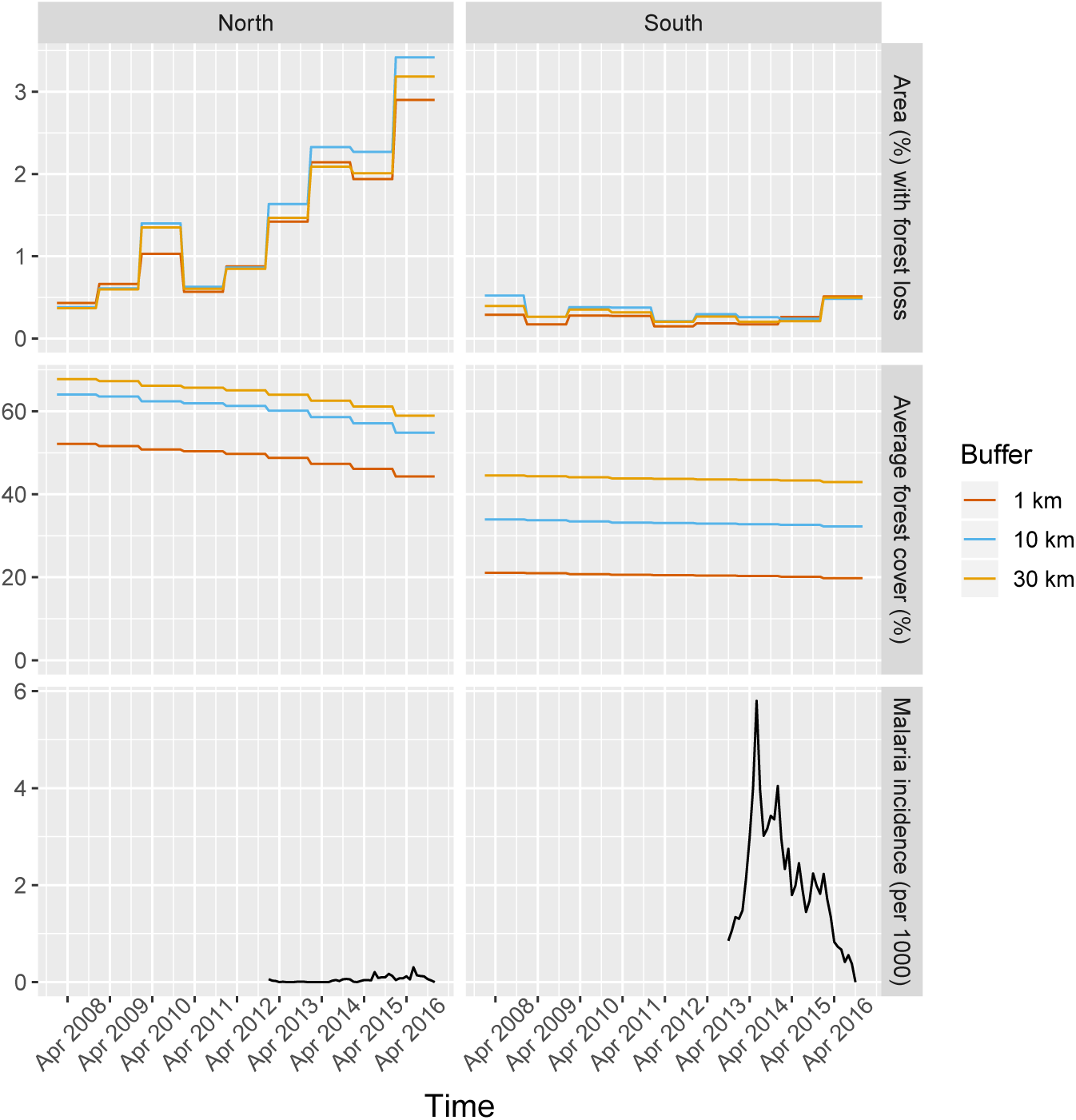
Time series of malaria incidence, forest cover (average tree crow cover around villages) and deforestation (percent area that experienced forest loss around villages), averaged over study’s villages and for varying buffer radius around villages (1, 10 and 30 km).

**Figure 21.**
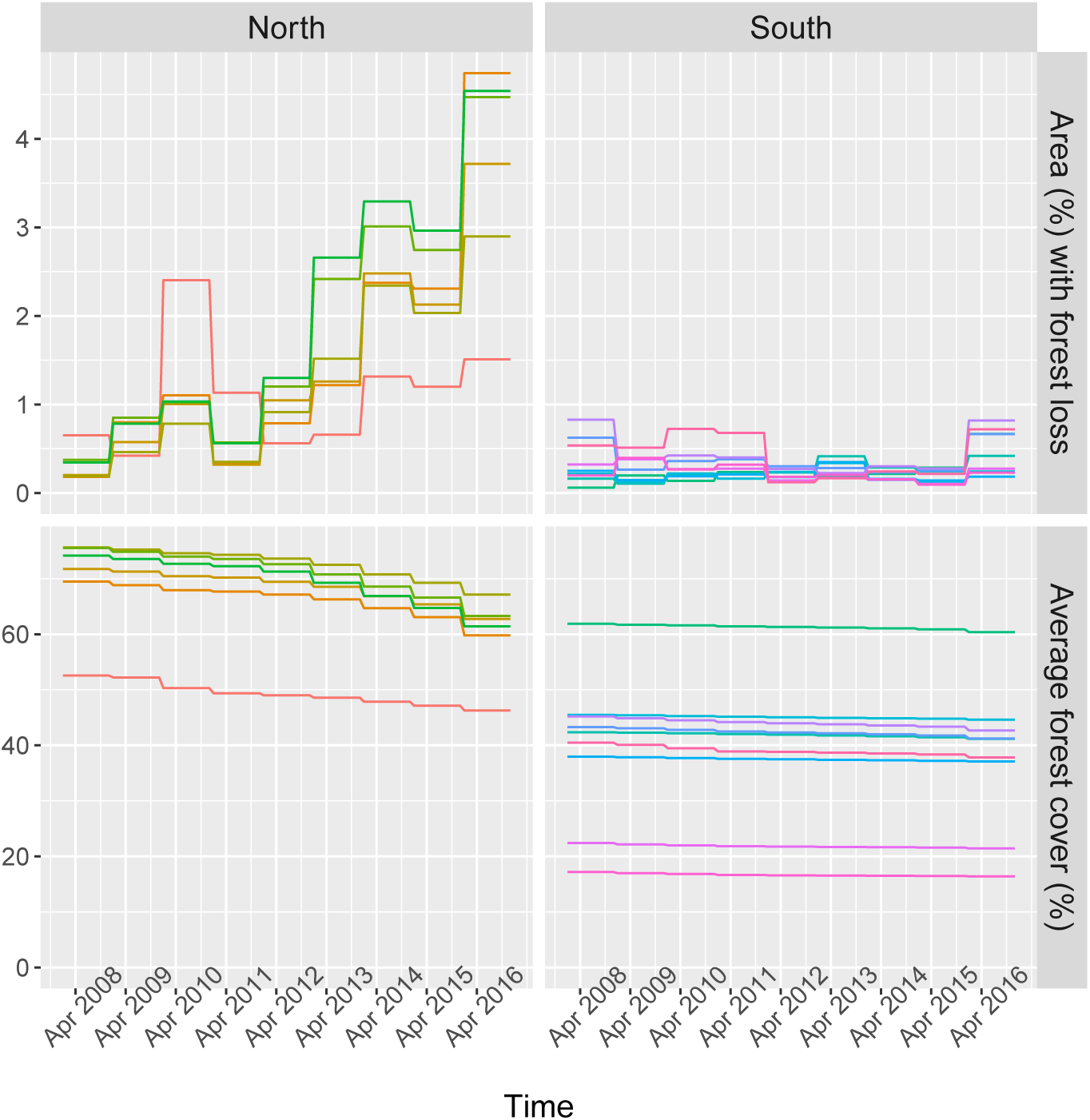
Time series of forest cover (average tree crow cover within 30 km of villages) and deforestation (percent area that experienced forest loss within 30 km of villages), for a few randomly sampled study’s villages. Each color represents 1 village.

**Figure 22.**
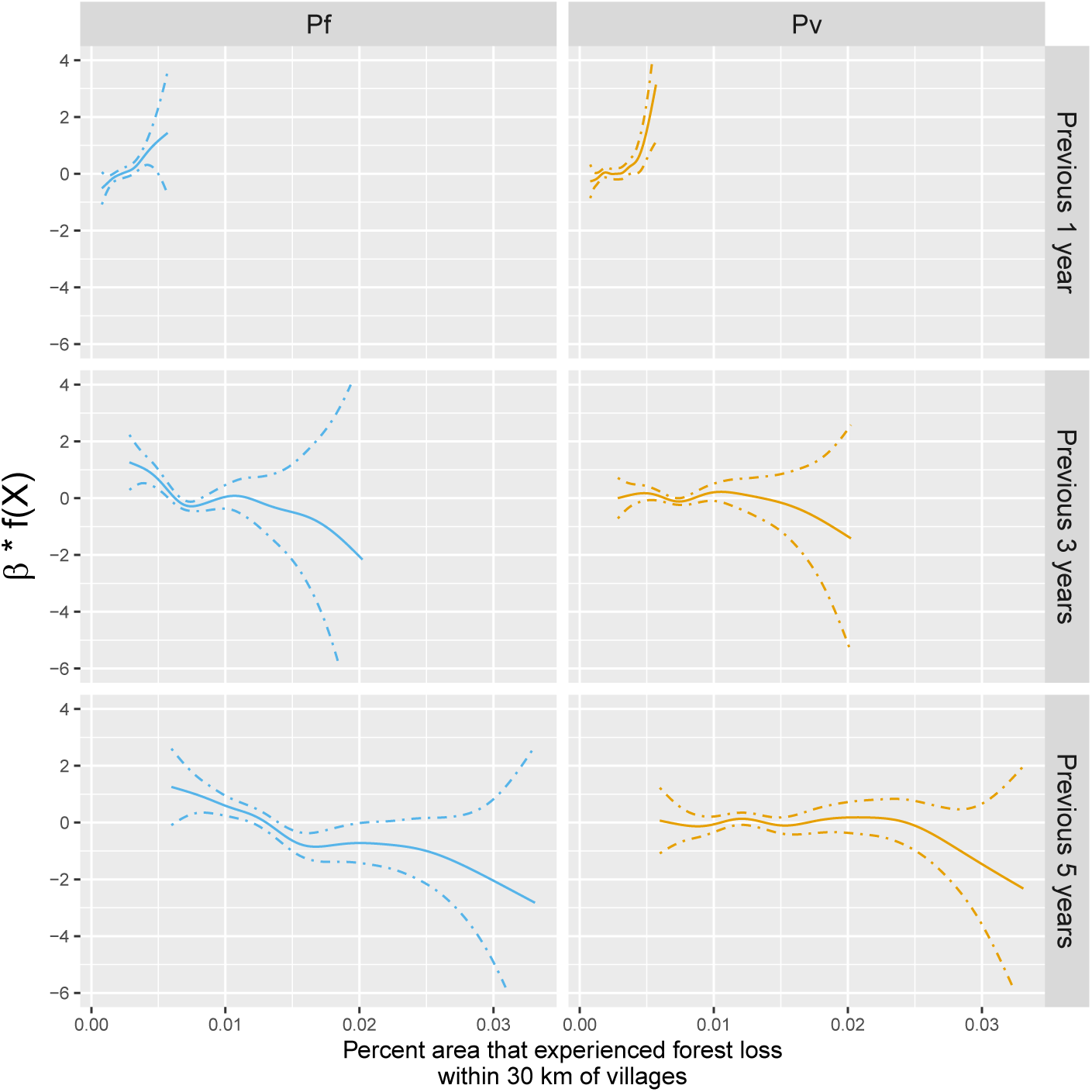
Adjusted relationship between deforestation and species-specific malaria incidence n southern Lao PDR. All models were adjusted for environmental covariates and forest cover selected by the model selection approach as described in the methods section on top of the spatio-temporal structure of the data (*f* (*t*), *f (Lat, Long)* and village random intercepts).

### S6: Additional figures

This section presents additional figures mentioned in the text and in the additional results section of the supplemental materials.

